# Climate warming is expanding dengue burden in the Americas and Asia

**DOI:** 10.1101/2024.01.08.24301015

**Authors:** Marissa L. Childs, Kelsey Lyberger, Mallory Harris, Marshall Burke, Erin A. Mordecai

## Abstract

Climate change is expected to pose significant threats to public health, particularly including vector-borne diseases. Despite dramatic recent increases in the burden of dengue that many anecdotally connect with climate change, the effect of past and future anthropogenic climate change on dengue remains poorly quantified. To assess the link between climate warming and dengue we assembled a dataset covering 21 countries in Asia and the Americas, and found a nonlinear relationship between temperature and dengue incidence with the largest impact of warming at lower temperatures (below about 20°C), peak incidence at 27.8°C, and subsequent declines at higher temperatures. Using this inferred temperature response, we estimate that historical climate change has increased dengue incidence by 18% (11 - 27%) on average across our study countries, and that future warming could further increase it by 49% (16 - 136%) to 76% (27 - 239%) by mid-century for low or high emissions scenarios, respectively, with some cooler regions projected to see dengue doubling due to warming and other currently hot regions seeing no impact or even small declines. Under the highest emissions scenario, we estimate that 262 million people are currently living in places in these 21 countries where dengue incidence is expected to more than double due to climate change by mid-century. These insights highlight the major impacts of anthropogenic warming on dengue burden across most of its endemic range, providing a foundation for public health planning and the development of strategies to mitigate future risks due to climate change.

## Introduction

Anthropogenic climate change is a major health threat that is already causing significant morbidity, mortality, and economic loss through its effects on biological processes and ecological and economic systems ^1,2^. Describing the causal relationship between climate change and its impacts on humans and the environment is necessary to anticipate and respond to health hazards and to attribute harms to fossil fuel emissions as part of climate accountability and justice efforts^3–5^.

Although effects on vector-borne disease were one of the earliest-recognized potential health impacts of climate change, few comprehensive estimates or attribution studies estimate quantitative, causal effects of climate change on vector-borne diseases to project changes in disease burden. Most previous climate attribution and impact studies have focused on short-term all-cause mortality, which is unlikely to capture the lagged impacts of temperature on disease transmission ^5,6^. Yet, understanding climate-driven changes in infectious diseases is critical given their large burden distributed unequally across the globe—disproportionately affecting lowand middleincome countries and concentrated around the tropics—often predominantly affecting younger people ^7^, and their potential to respond differently to temperature changes than other direct heat impacts that are not mediated through transmission^8^.

The statistical techniques used to isolate climate impacts on health, which often rely on temporal variation within locations over time, require globally representative panel data. The lack of these data for most infectious diseases, in comparison to the vital statistics data and electronic health records frequently used for all-cause mortality estimates, has made it difficult to exploit causal approaches to climate attribution for infectious diseases. Instead, studies centered on estimating the temperature dependence of vector-borne diseases or projecting future climate impacts have primarily used process-based mechanistic models (e.g., refs^9–14^), leveraged spatial variation in disease incidence (e.g., refs ^15,16^), or studied a single location or region^17^ (although see refs ^18,19^). These studies face three main challenges. First, while mechanistic models can incorporate known biological processes, they cannot capture all mechanisms and complexities underlying the transmission process, resulting in relative estimates of suitability rather than absolute estimates of human cases or incidence ^17,20,21^. Second, given the strong spatial correlation between temperature, other weather variables, land cover, and social and economic conditions, concerns about omitted variable bias make isolating the causal effect of temperature challenging in cross-sectional studies ^22^. Finally, studies leveraging temporal variation in dengue incidence and temperature within a single region may not generalize to other areas.

Dengue in particular has emerged as a rapidly expanding infectious disease, causing an estimated 96-105 million symptomatic cases per year, concentrated primarily in Southeast and South Asia and Central and South America ^23,24^. Dengue, like most vector-borne diseases, is acutely sensitive to temperature due to its ectothermic vectors, and a large body of literature has advanced an understanding of vector thermal biology and climatic suitability for transmission. These studies suggest clear mechanisms and pathways by which temperature may impact dengue, with nonlinear impacts on suitability peaking between 26 and 29°C and impacts occurring 1-3 months after changes in temperature ^11,21,25^.

Here, we study the impact of temperature on dengue transmission in the Americas and Asia by integrating data from 21 dengue-endemic countries spanning a large temperature gradient and wide range of climatic conditions in order to estimate the temperature-dependence of dengue in-cidence and the role of climate change in current and future dengue burden (Fig. 1). These countries span the majority of high incidence endemic areas, although notable absences are India and Africa (Fig. S1). We use subnational, monthly reported cases that cover 6,639 administrative units to directly estimate the relationship between temperature and dengue incidence (Fig. S2, Fig. S3).

**Figure 1:**
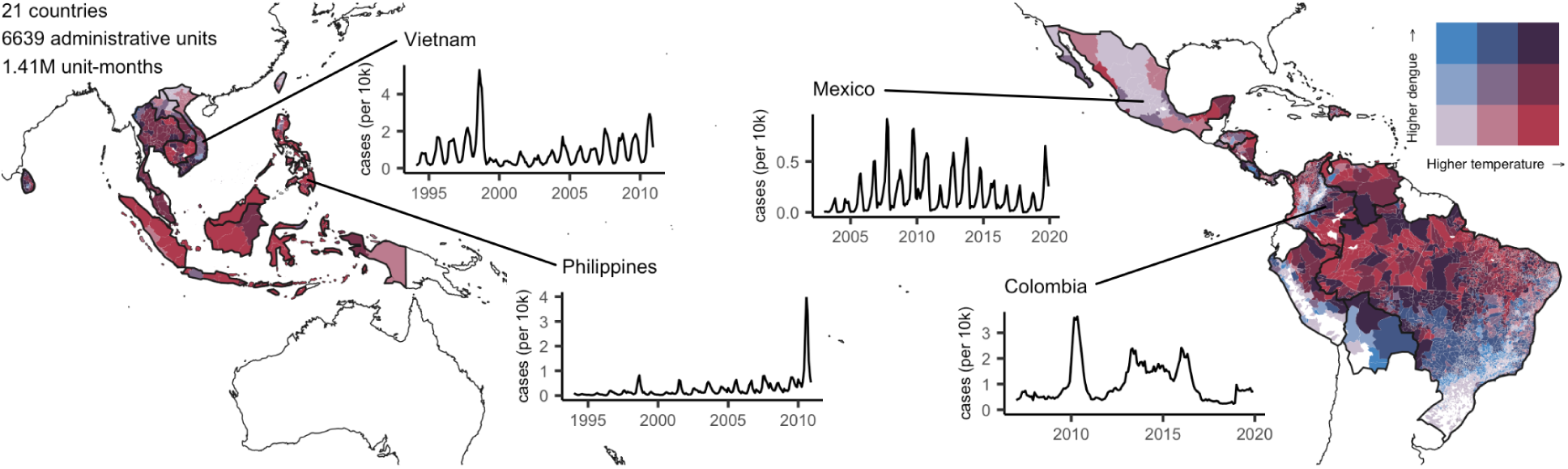
Subnational data on dengue and temperature from 21 countries. Darker red indicates warmer temperatures and darker blue indicates higher incidence of dengue. White areas within study countries do not have reported dengue cases in the database. Insets show four examples of epidemic dynamics in different countries.

We leverage the panel structure of the data with repeated observations for different administrative units to allow us to control for time-invariant, location-specific factors, regionally distinct seasonal patterns, and broad scale temporal patterns at the country level driven by factors other than temperature, like urbanization, invasion of new dengue serotypes, or changes in policy. In doing so, we estimate the temperature dependence of dengue based on anomalies in both temperature and dengue incidence. As deviations in temperature from the average conditions in a given space and time are plausibly random, we can interpret our result as a causal estimate. In contrast to models designed for prediction or explaining maximal variance in dengue incidence, we aim to isolate the specific effect of temperature in order to project relative changes in dengue from climate change while holding non-climatic factors fixed. We build on the biological understanding of temperature sensitivity of mosquito vectors and transmission established in previous studies of dengue thermal biology and geography, incorporate up to 4 months of temperature lags, and allow for a nonlinear relationship between dengue cases and daily temperatures (see Methods). Given the complex, nonlinear dynamics of disease transmission including the important role of population immunity, to confirm that this statistical approach will isolate the ex-pected temperature response in a complex dynamic system, we also simulate a mechanistic model of transmission with a known temperature relationship, then use our statistical approach on the simulated disease data to estimate the temperature response (see Methods). We find that this statistical approach accurately identifies the shape of the temperature response, particularly the temperature of peak transmission (Fig. S4).

We further test for potential sensitivity in the model to different modeling choices, including different lag structures and degrees of nonlinearity, different controls and fixed effects, and different weighting of spatial units in the dataset. Given the potential for the relationship between temperature and dengue to vary ^25,26^, we also examine whether covariates like average dengue incidence, continent, healthcare expenditure, population density, and previous dengue incidence at multiple lag times (which may capture some variation in population immunity and/or serotype interactions) significantly moderate the relationship between dengue and temperature. Using our estimated dengue-temperature relationship, which we find is robust to these potential sources of heterogeneity, we then project changes in dengue incidence due to historical and future anthropogenic warming using multiple future scenarios across a range of climate models to capture uncertainty. While we control for precipitation, as well as large-scale patterns that may affect entire countries, such as the El Niño Southern Oscillation (ENSO), when estimating the temperaturedependence of dengue, we focus solely on the impact of changes in average temperatures un-der climate change and do not project changes due to other potential impacts of climate change given their uncertainty.

## Results

### Thermal sensitivity of dengue incidence

We find that dengue incidence responds nonlinearly to temperature, increasing up to a peak at 27.8°C (95% CI: 26.9 - 29.4°C) and declining at higher temperatures (Fig. 2), consistent with mechanistic models ^11^. This dengue - temperature response implies that the effect of increasing temperature is largest at low temperatures (below about 20°C), declines to zero at 27.8 °C, and becomes negative at higher temperatures. These effects are large in magnitude, with a 1°C increase in constant temperature associated with approximately a doubling of dengue incidence in cool regions (15-20°C), consistent with a comparison of seasonal anomalies in dengue and temperature (Fig. S6). The general shape and nonlinearity of this relationship is similar across a range of alternative model specifications, including higher degree polynomials, varied lags of temperature, alternative fixed effects and weightings, and removing Brazil, which makes up 74% of the spatial units in our dataset (Fig. 2, Fig. S5, Table S1).

**Figure 2:**
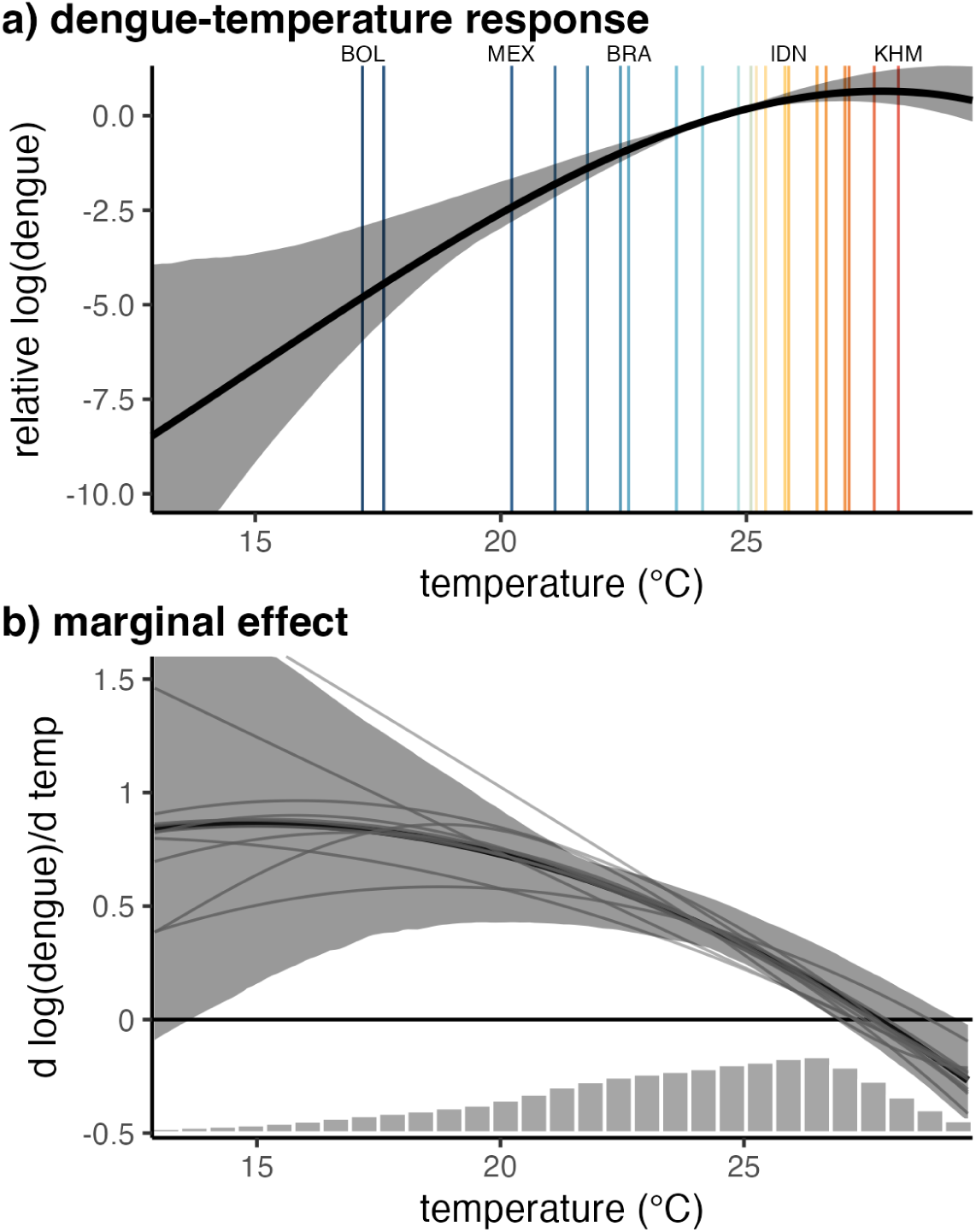
Effect of temperature on dengue. (a) Global nonlinear relationship between dengue cases and temperature and (b) the slope of that relationship indicating the marginal effect of temperature on dengue incidence. Main panel regression model fit in black with 95% confidence interval in gray shading. Vertical lines in (a) indicate country mean temperatures, with labels highlighting the coldest and warmest countries as well as the three highest population countries in the sample: Bolivia (BOL), Mexico (MEX), Brazil (BRA), Indonesia (IDN), and Cambodia (KHM). Thin gray lines in (b) represent variations on the main model using alternative specifications. Histogram in (b) shows the distribution of observed monthly temperatures. Model estimates are restricted to the 1st to 99th percentiles of the observed temperature distribution. We anchor the relative log(dengue) at 0 at the population-weighted average monthly temperature in the sample (24.5 °C). Central estimate for the main model is from the full sample, and confidence interval is calculated from bootstrapped regressions.

These results also align with previous mechanistic models based on laboratory-derived mosquito thermal performance curves, which predicted maximum dengue transmission at 29°C for *Ae. aegypti* and 26°C for *Ae. albopictus* ^11^. We estimate that precipitation also has an effect on dengue incidence, with some evidence of larger positive marginal effects at low precipitation levels and smaller impacts of at high rates of precipitation (Fig. S7).

The impacts of temperature changes on dengue incidence may depend on existing conditions, including local serotype dynamics, population immunity levels from previous exposure, availability of vector breeding habitat, the dominant vector species (with *Ae. aegypti* having a slightly warmer thermal optimum), living conditions and exposure to vectors, and public health responses. We focused on investigating whether the thermal sensitivity of dengue incidence varied spatially over large continental regions (e.g., Asia and the Americas), with country health expenditure, with population density, and with average dengue incidence. Importantly, we cannot causally attribute variation in estimated effects to any of these covariates due to the spatial correlation with many other factors that may drive variation in the impacts of temperature.

None of these potential sources of heterogeneity had significantly different incidence - temperature relationships, but the largest variation in thermal responses was between continental regions, with smaller variations by health expenditure, population density, and current dengue incidence (Fig. 3). For all responses, we see larger positive marginal effects at low temperatures, a decline to zero around 27-30°C, and negative marginal effects at higher temperatures. The temperature responses differed slightly in the exact optimum temperature and the magnitude of the marginal impacts at low temperatures. The temperature response for Asia displayed a slightly higher optimum temperature than the Americas, and smaller marginal impacts at low temperatures, although support in the lower temperature range was limited for Asia given the very few observations at low temperatures there. For all other factors we did not find strong qualitative or quantitative differences in the incidence - temperature relationship despite *a priori* hypotheses that factors like healthcare expenditures, population density, and average dengue incidence might be moderators. In particular, we did not find evidence that average dengue incidence modulated the incidence - temperature relationship, despite two competing hypotheses that high historical incidence could either reduce the temperature sensitivity of incidence (by depleting the pool of susceptible people and/or enhancing healthcare system preparedness) or increase it (by promoting serotype interactions that can cause symptomatic and severe dengue and/or by indicating environmental and socioeconomic conditions that are more conducive to dengue transmission). Before projecting estimated changes in dengue incidence under climate scenarios, we further sought to investigate whether the marginal effect of temperature differs by existing dengue incidence and susceptibility. While the previous analysis using average dengue incidence relied on spatial variation between locations, temporal variation in dengue incidence within locations could also result in differential impacts of temperature. In particular, decreasing population susceptibility (or increasing immunity) could lead to smaller impacts of temperature; in the extreme case, in a fully immune population, changes in temperature would have no impact on dengue incidence. To test for this, we estimated differential temperature responses by population susceptibility in both simulated and observed data. In simulated disease dynamics (see Methods), we found little difference in the marginal effect of temperature when interacting with 1-month-lagged population susceptibility categories (Fig. S8). Similarly, we took advantage of the long time span of our observed dengue data that captures a large range of existing dengue immunity levels to empirically investigate whether year-to-year variation in population susceptibility affected the temperature response of incidence. As we lacked information on population susceptibility in the observed data, we constructed proxies of population immunity using 1-year and 3-year moving windows of lagged cases–specifically 1-12 months, 1-36 months, 7-18 months, and 7-42 months– based on existing literature, which suggests that short-term immunity to all dengue serotypes after an initial infection wanes in 1-3 years^27–29^. We then interacted this population immunity measure with temperature in the panel regression (Fig. S9). We estimated remarkably similar effects of temperature on dengue incidence across different levels of the immunity proxies (Figure S10a). However, even small observed changes in responses could result in differential sensitivity to future changes in temperature depending on the distribution of temperature. To quantify the impact of the differences in responses, we calculated the marginal effect of temperature over the distribution of current temperatures (1995 - 2014) and we found only small differences in the average estimated marginal effect of temperature (Figure S10b). These minimal differences across a range of immunity levels suggest that at least based on historical dengue dynamics, we see little evidence that effects of temperature on dengue will decline due to decreasing susceptibility. These findings are consistent with waning immunity and the invasion of new strains and serotypes, which prevent populations from reaching persistent herd immunity to dengue.

**Figure 3:**
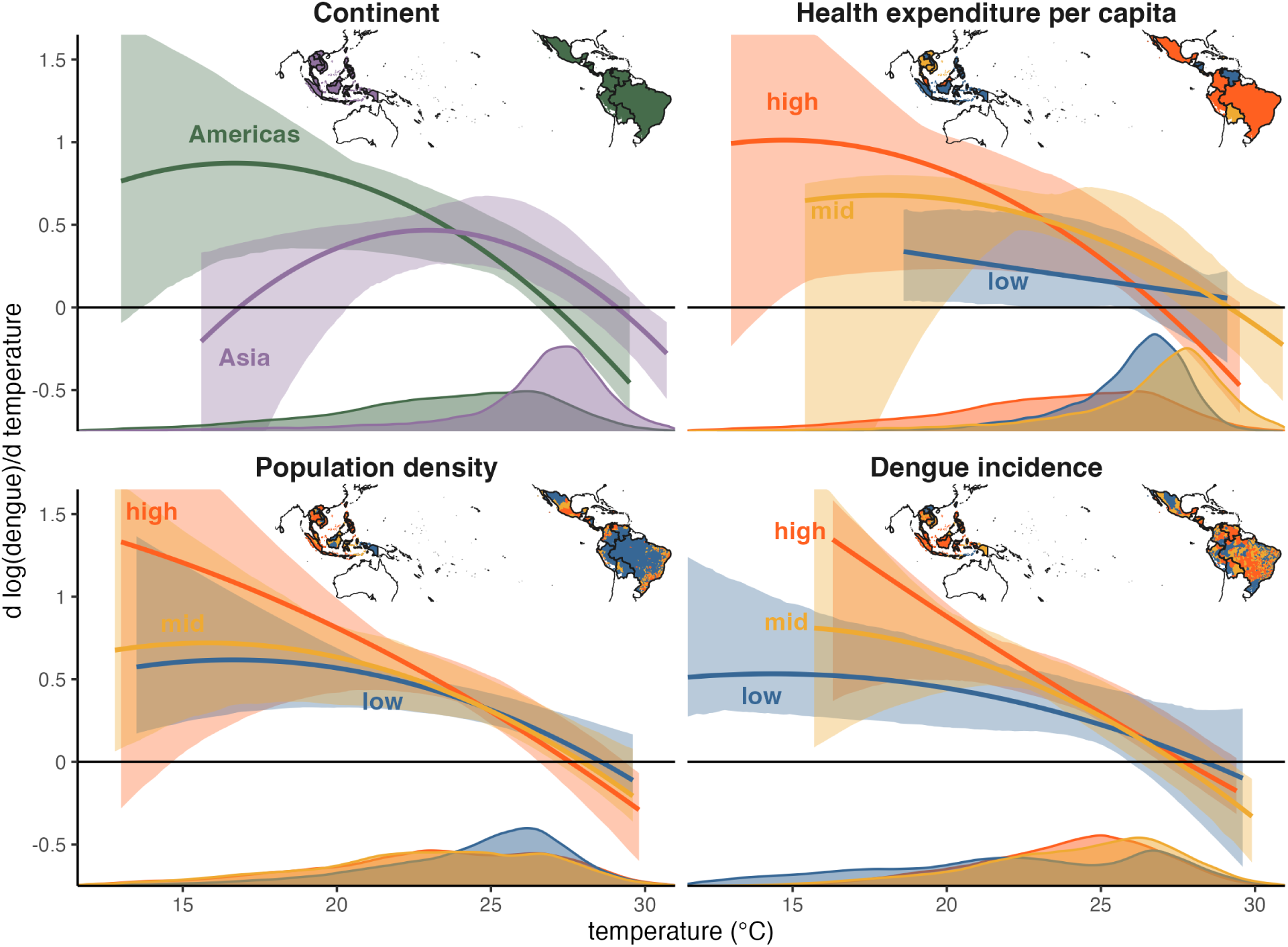
Spatial heterogeneity in the dengue-temperature relationship. Marginal effect of temperature on dengue incidence by (a) continental region, (b) health expenditure tercile, (c) population density tercile, and (d) dengue incidence tercile. Dengue incidence and population density are subnational covariates, and continent and health expenditure are country-level covariates. Lines are the mean estimates and shaded areas are 95% confidence intervals, with estimated marginal effects trimmed to the 1st to 99th percentiles of the observed temperature distribution for each tercile or continental region. Where relevant, confidence intervals were truncated for visibility. Density plots show the distribution of monthly temperatures for each tercile or continental region and inset maps depict the spatial units included in each tercile for the different covariates, with colors matching those of the estimated marginal effects in each panel.

### Historical and future impacts of climate change

To quantify the impact of historical climate change, we estimated the change in dengue incidence under observed temperatures for 1995-2014 compared to a counterfactual of global circulation models (GCMs) without anthropogenic forcing. The GCM ensemble suggests temperatures were 0.90°C higher on average across our study region in 1995-2014 due to anthropogenic emissions since the preindustrial period, with variation among locations (Fig. S11). Incorporating these locally-specific climate change estimates into our inferred temperature - dengue incidence function, we estimate that on average over the locations included in the study, 18% (95% CI: 11 - 27%) of current dengue incidence is due to this anthropogenic climate warming that has already occurred (Fig. 4, Table S2). The estimates vary both within and between countries, with cooler areas in Bolivia, Peru, Mexico, Brazil, and Colombia seeing 30-40% of existing dengue from climate change and warmer countries like Thailand and Cambodia having little impact from historical warming. Most of the variation in these effects is driven by differences in baseline average temperature as the differences in temperature change due to historical climate warming between locations are small (Fig S11).

**Figure 4:**
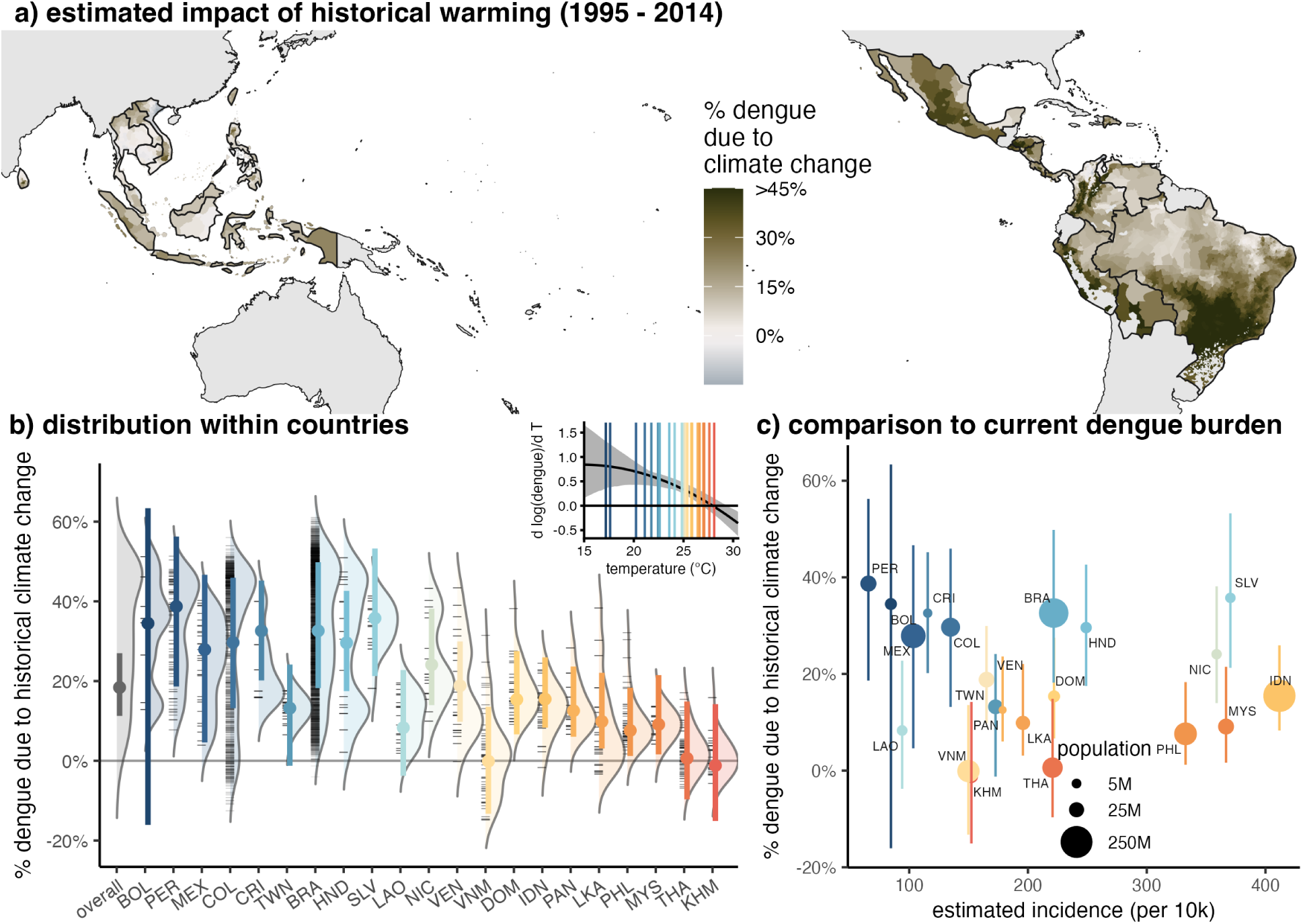
Climate change has already increased dengue incidence. (a) Estimated percent of dengue burden attributable to anthropogenic climate warming for administrative units with observed dengue cases during the study period, estimated as the average % change in dengue between observed and counterfactual climate during 1995-2014. (b) Distributions show variation across administrative units within each country. Black tick marks indicate individual unit mean values, and colored points and bars show the mean and 95% CI of the population-weighted average estimate by country. Countries are ordered by current average temperatures with warmer countries to the right. Only administrative units with reported dengue are included in the distributions and country averages. Inset: marginal effect of temperature on dengue from the main model specification, with country-average temperatures indicated with vertical lines matching the colors in the main panel. (c) Impacts of historical warming are largest in cooler countries where current incidence is low, but impacts are also substantial in moderate temperature countries with high current dengue burdens (e.g., Brazil, Honduras, El Salvador, and Nicaragua). Point colors (indicating temperature) are the same in (a) and (b) and point sizes in (c) indicate population size. Line ranges are 95% CIs as in (b).

These impacts of climate warming equate to a large number of dengue cases and affected people. While some of these cooler countries where a larger portion of existing dengue is due to historical climate change have relatively low current dengue burdens (e.g., Peru, Bolivia, and Mexico), other countries like Brazil, El Salvador, and Indonesia with intermediate average temperatures (22-26°C) have high current burdens of dengue and 10-30% of that existing burden is estimated to be due to climate change (Fig. 4). Using the empirical dengue incidences in the dataset and 2015 population estimates, this translates to 4.3 million cases per year (2.8 - 6.2 million cases) due to climate warming in the study countries. When accounting for underreporting using estimates of dengue burden ^23^, these estimates are ten times larger: 46.1 million (29.1 - 73.5 million) cases each year due to anthropogenic climate change that has already occurred in the 21 study countries.

Given the strong and robust relationship between temperature and incidence, future warming will also increase dengue incidence. On average in the study region, we predict a 76% (95% CI: 27 - 239%) increase in dengue incidence under the high emissions scenario (SSP3-7.0) by midcentury (Fig. 5, Table S2). Under the low emissions scenario (SSP1-2.6), we estimate a smaller but still substantial 48% (16 - 136%) increase in dengue incidence. Our analysis of the intermediate emissions scenario (SSP2-4.5), detailed in the supplementary material, falls between the estimates for SSP1-2.6 and SSP3-7.0. While some low-elevation equatorial areas will warm beyond the optimal temperature for dengue transmission and are projected to see small declines in dengue incidence due to warming, the majority of locations are projected to see increases in dengue incidence under all emissions scenarios (Fig. 5, Fig. S12). Some cooler regions of Mexico, Peru, Bolivia, and Brazil are predicted to see over 150% increases in dengue incidence due to climate warming under all emissions scenarios. Estimates in Bolivia in particular are very uncertain under the high emissions scenario, with population-weighted average dengue projected to increase by 811.1%(95% CI: 22.4 - 6101.4%). Many of the largest cities in the Americas are located in these cooler regions where large increases in dengue are projected (Fig. 5, black circles), and 262 million people in these 21 countries are currently living in places expected to see over 100% increases in dengue incidence under the high emissions scenario by mid-century (based on 2015 population and mean estimates in each location). Among the 21 countries included in the study, 15 are projected to see increases under all emissions scenarios. Based on current dengue incidence and population in the 21 study countries, these effects translate to an additional 68.0 million infections (28.7 - 136.1 million; 7.3 million [3.1 - 14.5 million] without rescaling for underreporting) under the low emissions scenario or 96.0 million infections (44.4 - 185.8 million; 10.9 million [4.4 - 21.4million]) under the high emissions scenario, compared to the estimated 4.3 million warming-driven cases per year (2.8 - 6.2 million cases) that have already occurred during the 1995-2014 period. Moreover, these future incidence numbers do not account for potential changes in population growth and dengue incidence due to non-temperature impacts in the future.

**Figure 5:**
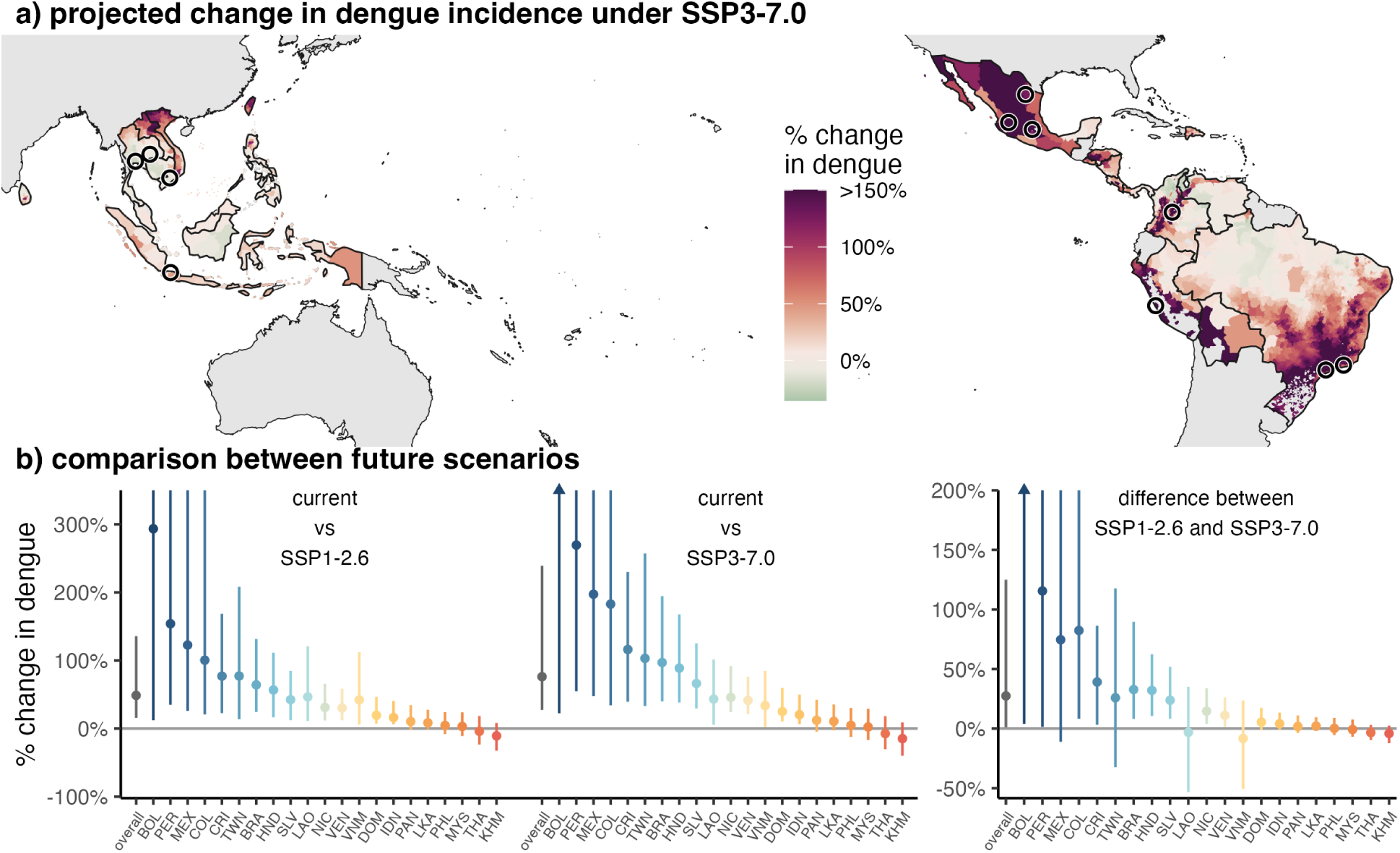
Estimated impacts of climate change on dengue incidence for 2040-2059 are widespread and largest in more temperate regions. (a) Most locations are expected to see an increase in dengue under climate change, with a small fraction of locations projected to experience slight declines due to temperatures exceeding the inferred optimal temperature. Many of the areas where large increases are predicted (shown in dark red), especially in the Americas, are areas with large cities and high population density. Black circles show cities over 5 million in population ^45^. Administrative units are colored by the mean percent change in dengue incidence under the high emissions scenario at mid-century (SSP3-7.0 in 2040-2059) compared to current temperatures (1995-2014). (b) Across all future climate scenarios, dengue incidence is predicted to increase for a majority of countries, with the largest increases in cold countries (left panel), and these impacts are estimated to be up to 100% larger for some countries under the highest emissions scenarios compared to the lowest (right panel). Estimated impacts are in percent change from current incidence in comparison between current and future scenarios, and the absolute difference between percentage changes in comparison between between different future scenarios. Countries retain ordering by average temperature, and country colors are consistent across all figures. Points show mean estimates and lines show 95% CIs. Points and confidence intervals are limited to 300% in comparisons between current and future and to 200% in comparisons between future scenarios for visibility. Where mean estimates are limited, points are shown as triangles.

Despite the estimated increases in dengue in the majority of locations even under the most optimistic (low emissions) scenario (SSP1-2.6, Fig. S12), we find a substantial benefit of mitigating carbon emissions for reducing projected future dengue incidence. We project that there would be an additional 27% increase (95%CI: 1 - 125%) under the high emissions scenario (SSP3-7.0), with some countries seeing additional 50-100% increases in dengue incidence, compared to SSP1-2.6 (Fig. 5b, Table S2). These estimates are largely similar when using continent-specific temperature responses, which showed the greatest difference between temperature responses, with the largest differences in projected impacts in Asia where no countries are predicted to have see significant declines in dengue under future climate scenarios due to the slightly higher estimated temperature where dengue incidence peaks (Fig. S13).

## Discussion

Despite substantial understanding of dengue vector thermal biology ^11,21,26,30^, the precise relationship between dengue incidence and temperature had not previously been quantified^17,25,31,32^. Yet, understanding this relationship precisely is critical, both for projecting impacts under climate change to better anticipate future changes and design adaptations to meet them, and for attributing changes in dengue to anthropogenic climate change that has already occurred.

We project that climate warming will increase dengue incidence in most areas, in contrast to malaria, which is projected to decline in sub-Saharan Africa by mid-century due to climate change-driven temperature increases ^19^. These divergent impacts are consistent with the differences in thermal biology between malaria and dengue, with malaria predicted to have a cooler optimum temperature around 25°C compared to the 27.8°C optimum inferred here and the previous mechanistically predicted 26-29°C optimum for dengue ^13,33^. Impacts of climate warming on dengue differ from to other health impacts of climate change, especially direct heat-related mortality, which is predicted to increase the most in already warm regions^6^, in that they are projected to be largest in relatively cool endemic regions, while a few of the hottest regions in our study are projected to see declines in dengue incidence due to climate change (Fig. 5). In fact, our study likely underestimates these future warming-driven dengue increases in cool areas as many such regions do not yet have consistent dengue transmission and reporting, and thus are not included in our dataset.

While our results highlight the benefit of climate mitigation in reducing the projected increases in dengue incidence, the projected increases under all scenarios suggest that climate adaptation that accounts for the public health consequences of dengue expansions will also be necessary even in the best case ^34^ (Fig. 5). Moreover, the consistency of the estimated dengue-temperature relationship across places with both high health expenditures and high current dengue incidence (Fig. 3) suggests that even the locations most likely to have better existing public health systems and dengue control programs will be strongly affected.

Although this work presents the most globally comprehensive (spanning 21 countries) quantitative estimate of temperature impacts on dengue to date, our estimates are constrained by the set of locations with available sub-annual, sub-national dengue data, which are primarily located in tropical areas of the Americas and southeast Asia. The study omits both other tropical areas with endemic dengue (notably south Asia and sub-Saharan Africa, Fig. S1) and cooler temperate regions that do not have consistent dengue transmission. As a result of the latter, our estimates focus on the impact of temperature changes on intensifying dengue incidence in endemic areas rather than expansion of dengue into new regions. Taken together, this suggests that our estimated change in dengue cases from climate change will be conservative due to the omitted dengue-endemic regions and the temperate regions where dengue transmission occurs sporadically and could expand ^35^ (including recently in California, Texas, Hawaii, and Florida, USA^36^ and in many European countries^37^). While some of the excluded areas may be at or above the optimum temperature for dengue transmission and see slight declines in dengue incidence due to climate change, the majority will see increases with future warming if their thermal responses are consistent with the study countries (Fig. S1). The estimated impacts of historical warming are also conservative as they use simulated temperatures without anthropogenic emissions only through 2014 (from the most recent Coupled Model Intercomparison Project Phase – CMIP6), and additional warming has occurred since then.

In addition to focusing on endemic regions, our projections center on the effect of changes in average temperature rather than any of the other impacts caused by climate change, including altered precipitation patterns, changes in temperature variability, increased extreme weather events ^38,39^, and behavioral adaptation to climate change. Further, our temperature response is estimated from real dengue dynamics in the field, and in doing so, implicitly incorporates complexities like serotype dynamics, vector control, policy, and population growth and mobility. Our projections assume that these other factors will not change in a way that alters the estimated dengue - temperature response (i.e., in a way that is interactive with temperature), an assumption largely consistent with the minimal differences in temperature impacts across a range of population susceptibility levels and socio-demographic contexts. Finally, our projections focus solely on the impact of temperature on dengue incidence, and the landscape of dengue transmission in the future will also depend on a range of other factors including urbanization, migration, the emergence of new serotypes ^40^, and/or potential medical advances in treating or preventing dengue ^41^, each of which could modify future temperature effects. While our model controls for precipitation, we do not estimate potential impacts of changing patterns of precipitation as these projections have high uncertainty^42^. Future work could build on this study to consider both precipitation and temperature to understand which climatic conditions are most constraining in different locations, and where projected changes in each is likely to have larger effects. Another important extension of this work is to estimate the burden and future trends in warmingattributable dengue in countries not included in the currently available 21-country dataset, including in South Asia and sub-Saharan Africa.

Our study contributes in several important ways to the growing body of literature attributing climate impacts on health ^5^. First, it makes quantitative predictions for how dengue burden will change under climate warming at the local, sub-national scale across a major swath of its endemic range, which can be used to develop targeted public health planning and responses and compare the consequences of different emissions scenarios. Second, this work provides a rare demonstration that theoretical models based on laboratory experiments can capture the thermal biology of complex infectious disease systems, reinforcing the idea that such thermal biology models can be used to predict climate responses in places with limited existing data. Third, it is among the first studies to attribute changes in infectious disease to historical climate change— expanding on the attribution literature centered on more direct effects such as heat waves, storms, and fires and providing a road map for future studies on other ecological and health impacts^19,43,44^. Finally, attribution studies like this one are increasingly used in litigation aimed at holding governments and fossil fuel companies financially accountable for negative societal effects of carbon emissions due to climate change, and developing funds to assist the most impacted countries for loss and damages.

## Methods

### Dengue case data

To obtain a comprehensive global dataset of dengue cases at the sub-annual and sub-national scale, we searched the Pan-American Health Organization (PAHO) and Project Tycho databases, Ministry of Health websites, and published literature. For Project Tycho, we selected “Dengue” for condition, “Month” as the interval type, and downloaded data for any country with *>*2 years of data at admin level 1 (state or province) or below. For all countries with endemic cases listed on the WHO website, we found their Ministry of Health (or equivalent) website and navigated to any relevant tabs or databases reporting health data. If we still could not locate relevant data, we searched the following on Google: “[country name] immunological bulletin OR dengue”. Additionally, we searched the literature using the same query for references to data sources; however, these were typically not publicly available. We found 25 datasets from 21 countries that span an average of 11 years (Table S3). We excluded datasets that spanned less than 2 years and trimmed our data to the end of 2019 to avoid confounding effects of COVID-19. We aggregated weekly data to monthly as follows: if a week spanned two months we split the cases into months based on the number of days within that week that fell into each month. To obtain incidence from the raw case counts, population size was obtained by summing population count within each administrative boundary (see “Subnational boundaries”) in the midyear of the time series, using population counts from WorldPop ^1^ on available on Google Earth Engine ^2^. In total, our dataset consisted of over 1.4 million monthly observations of dengue incidence at the first or second administrative level.

### Subnational boundaries

We matched the names of the subnational administrative units with those in shapefiles downloaded from the Humanitarian Data Exchange ^3^ except for Taiwan, which was downloaded separately^4^. Because case data in Costa Rica is reported for socioeconomic regions, which does not fall cleanly in administrative level 1 or 2, a shapefile of administrative boundaries within Costa Rica was manually modified to correspond to socioeconomic regions. We used the district (administrative level 3) shapefile and aggregated to six socioeconomic regions based on a mapping from canton (administrative level 2) to socioeconomic region^5^. Four districts (admin level 3) in socioeconomic regions different from the rest of their cantons are switched to the correct socioeconomic region.

### Historical temperature data

To obtain downscaled daily temperature values with consistent performance across regions, we compared ERA5 daily climate reanalysis product ^6^ to Global Historical Climatology Network (GHCN) station observations in the countries included in this study. We found that ERA5 temperatures were on average downward biased relative to GHCN station observations, with large (up to 10°C) negative biases at high elevations (Fig. S14). To avoid this differential bias in the ERA5 product, we debiased using WorldClim, a high resolution climatology product with monthly average temperature from 1970 - 2000^7^:

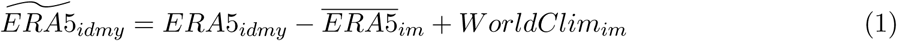

where 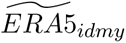 is the debiased daily temperature, *ERA*5*_idmy_* is observed ERA5 temperature in location *i* on day *d* in month *m* and year *y*, 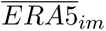 is the monthand locationspecific ERA5 climatology over 1970 - 2000 and *WorldClim_im_* is the WorldClim climatology over the same time period. We calculated 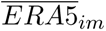 from monthly average temperature in ERA5 monthly products, and used *ERA*5*_idmy_* from the daily average temperature product available on Google Earth Engine ^2^. We found that this correction reduced the mean error between satellite-based temperature estimates and ground-based measurements from the GHCN, especially at higher elevation sites (Fig. S14). We calculated higher powers of debiased temperature data (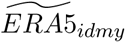) for each day and grid cell to allow for potential nonlinearity in the temperature-dengue relationship at the finest spatial and temporal scale, then calculated administrative unit - month populationweighted averages using the subnational boundaries described above and WorldPop population estimates ^1^. We used the same population-weighted average approach to calculate monthly average precipitation from ERA5.

### Climate scenario temperature projections

To estimate the change in dengue transmission under future climate change, we used projected temperature from the Coupled Model Intercomparison Project Phase 6 (CMIP6)^8,9^ under different scenarios. In keeping with recommendations from the latest IPCC report ^10^ and Hausfather et al. ^11^, we used SSP3-7.0 as a high baseline emissions scenario, and SSP2-4.5 and SSP1-2.6 as medium and low emissions scenarios, respectively. We also considered the historical-natural projections as our counterfactual for historical temperature absent anthropogenic forcing to understand how climate change has already impacted dengue transmission. Of the 39 global climate models (GCMs) available from the CMIP6, we followed recent guidance on excluding “hot models” and limit to models with transient climate response (TCR) in the likely range (1.4 - 2.2°C)^11^ using existing TCR calculations for the GCMs^12^. We further limited to models that have monthly temperature available for projections for all three future climate scenarios specified above, resulting in 21 GCMs included in this analysis. A full list of included models and scenarios can be found in Supplementary Table S4.

Given known biases in average temperatures and unreliable daily temperature anomalies in GCMs ^13,14^ we used the delta change method ^15^ to calculate temperature under future climate scenarios as follows. We determined the estimated change in temperature from the current period to the mid 21st century as

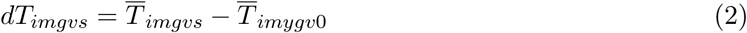

for location *i*, month of the year *m*, model *g*, variant *v* and scenario *s*, where 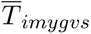 is the average of monthly temperatures within a specified period and 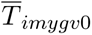 is the average from the historical scenario for 1995 - 2014. We performed this delta change method for both future and historical-natural scenarios , using the 2040 - 2059 time period for future scenarios and 1995 - 2014 for the historical-natural scenario when calculating 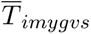. All scenarios used 1995 - 2014 for 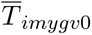, with the difference from the historical-natural scenario being the historical climate change and the difference from future scenarios being the potential climate change by mid-century. We matched variants between scenarios for each GCM when calculating *dT* , defaulting to the first variant available for all of the desired scenarios run for a GCM, or if no variant was available for all scenarios, a single variant for future scenarios and a separate variant for the historicalnatural scenario. Details on variants used for each GCM and scenario are in Supplementary Table S4. To calculate estimated temperature under different scenarios, we then added the estimated temperature change to debiased daily temperature data 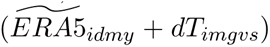, and then aggregated to monthly temperatures as described above. To compare scenarios, we used observed temperatures from 1995 - 2014 for all locations for the current period.

### Data extraction for moderators

For each country, health expenditure per capita expressed in international dollars at purchasing power parity in 2010 from WHO Global Health Expenditure Database was downloaded from World Bank Open Data ^16^. Taiwan was the only location in our dataset missing health expenditure information. For the Gini index and health expenditure, we selected the year that is the average of all time series midpoints (2010). If this was unavailable, we took health expenditure from the closest date. We calculated population density using total population from WorldPop^1^ (as described above) and dividing by the area of the administrative unit. To estimate average dengue incidence at the administrative units, we calculated annual average dengue incidence in the our case data, and scaled by the ratio between observed country-level average annual dengue incidence in the case data and country-level incidence estimates in 2017 from the Global Burden of Diseases ^17^ to account for potential differences in disease detection rates between countries while still allowing for variation in dengue incidence within countries.

### Estimating dengue-temperature responses

To obtain an overall estimate of temperature dependence on dengue we used a panel regression. Our model takes the form

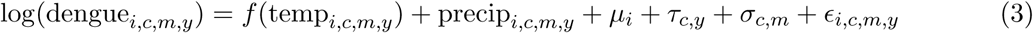

where *i* is unit, *c* is country, *m* is month, and *y* is year. This model includes an administrative unit fixed effect to control for differences between locations that are consistent over time, a countryyear fixed effect to account for country-level patterns over time, and country-month fixed effect to remove seasonal patterns in dengue and temperature. The main specification included population weighting and used cubic polynomials of temperature with 1-3 months of lags, but we also considered functional forms with up to degree 5 and lags from 0 to 4 months. We also tested sensitivity of model estimates to including quadratic effects of precipitation rather than linear, no weighting of estimates, removing Brazil from the sample, fixed effects for country - month - year rather than country - month and country - year (to account for potential interannual variation in seasonality at the country scale), and fixed effects for unit - month rather than country - month (Fig. S5). All models were run as Poisson regressions with the ‘fixest’ package in R^18^. For countries that had data from multiple sources, we removed any duplicate years from the earlier data source and treated each country-data source as a different “country” for the purpose of fixed effects to allow for potentially different reporting rates between data sources.

To understand how this relationship varies spatially, we interacted the estimated temperature relationship with administrative unit-level covariates including continental region, health expenditure, population density, average dengue incidence:

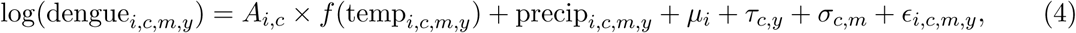

where *A_i,c_* is the covariate value for unit *i* in country *c*. For continent we divided the data into Asia and the Americas and for the remaining covariates, we split the data into terciles with health expenditure terciles being defined at the country-level due to availability of health expenditure data, and population density and dengue incidence terciles defined at the administrative unitlevel. We defined these terciles only accounting for administrative units that reported dengue during the study.

Additionally, population susceptibility levels may modulate the magnitude of the impact of temperature. Understanding whether temperature-sensitivity differs by population immunity levels is critical for future climate projections, as changes in temperature could lead to altered transmission rates, which in turn would impact population immunity and lead to differential subsequent temperature impacts. To quantify whether temperature impacts may differ by population susceptibility, we used temporal variation in population susceptibility in historical data given the long time series of data available with a range of observed dengue dynamics. This approach relies on the existing variation in dengue dynamics, which includes some locations with both unstable, highly episodic dynamics—consistent with multiyear cycles of outbreaks and higher population susceptibility—and other locations with steady seasonal dynamics—consistent with lower population susceptibility. While population susceptibility data is not available, we used lagged incidence as a proxy measure. In particular, we rescaled observed incidence using the countryspecific Global Burden of Disease ratios ^17^, and then calculated the average rescaled incidence over specified windows. Consistent with the existing literature on dengue immunity, which suggests cross-protection from all serotypes for 1-3 years after an initial infection followed by waning immunity^19–21^, we considered four different windows: 1-12 months lagged, 1-36 months lagged, 7-18 months lagged, and 7-42 months lagged, with the lagged windows beginning 7 months prior intended to exclude dengue incidence in recent months. Recent incidence could reflect the number of newly infected individuals, a measure of current transmission, rather than baseline population susceptibility levels. This allowed us to estimate a dengue immunity proxy, which we then binned into four quantiles (Fig. S9). We then interacted this time-varying immunity proxy with temperature as follows to estimate a temperature response for each quantile of dengue immunity (S10):

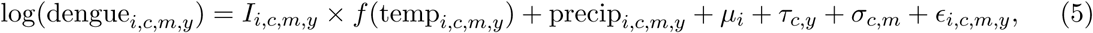

where *I_i,c,m,y_* is the immunity quantile for unit *i* in country *c* at month *m* in year *y*. This was estimated for all four lagged time windows that we considered.

We calculated confidence intervals on estimated temperature responses and marginal effects using block bootstraps and analytic confidence intervals based on the Delta method. Block bootstraps were conducted using countries as the blocks, and re-sampling entire countries with replacement, then using the full time series for each administrative unit in the country in the bootstrap sample. These block bootstraps served to preserve both the temporal correlations within locations as well as the spatial correlation within countries. Bootstrapped confidence intervals were slightly wider than analytic confidence intervals calculated with the delta method using standard errors clustered at the country-level (Fig. S5). Unless otherwise noted, all reported confidence intervals are based on block bootstraps.

To test whether this approach will isolate the impact of temperature in a dynamical infectious disease system, we simulated transmission in a mechanistic model with a known temperature response and showed that the statistical approach accurately identifies the shape of the temperature response (see Supplemental Methods).

### Attributing dengue burden and projecting future impacts

Using the estimated temperature relationship *f* (temp*_i,c,m,y_* ), we calculated the change in dengue incidence due to temperature changes as

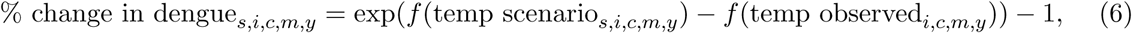

where *s* indexes the scenario of interest (SSP1-2.6, SSP2-4.5, SSP3-7.0, historical-natural forcing). We used debiased monthly ERA5 data for observed temperatures and monthly climate projections (described above) for scenario temperatures. We estimated the percent change in dengue for the specified GCMs (21 for future scenarios, 10 for historical - natural forcing, see Table S4) using 100 of the bootstrapped estimated temperature responses to incorporate uncertainty from both the model estimates and the climate scenarios. For each GCM and bootstrap, we calculated the average percent change in dengue over the 20 years of temperature data for each administrative unit. To estimate administrative unit-specific effects, we then calculated means and 95% confidence intervals over 2100 estimates from the GCMs and bootstraps. Similarly, to estimate country-level and overall effects, for each bootstrap and GCM we calculated a populationweighted average of the administrative unit averages using only administrative units with observed dengue cases in the dataset, then calculated the mean and 95% CI over the 2100 estimates for each scenario. We translated the estimated percentage changes into cases by multiplying the average percent change in dengue for each unit by dengue incidence and population, then summing over all units for each bootstrap and GCM, and similarly took the mean and 95% CI across the different bootstraps and GCMs. We estimated with both average annual dengue incidence in the sample for each administrative unit and incidence scaled by the Global Burden of Disease country estimates (see above in “Data extraction for moderators”). We used 2015 population estimate from WorldPop ^1^ for case estimates and population-weighted country/overall averages. To compare the between two future climate scenarios (particularly between SSP1-2.6 and SSP3-7.0), we used the scenario temperatures in equation 6 instead of observed temperatures. We projected future impacts using both the main model bootstrapped estimates and the continent-specific estimates.

## Acknowledgements

Some of the computing for this project was performed on the Sherlock cluster, and we would like to thank Stanford University and the Stanford Research Computing Center for providing computational resources and support that contributed to these research results. We would also like to acknowledge computational resources from Google Cloud for Earth Engine and thank Michael Sherman for his assistance applying for additional batch task quota through the uplift program. We acknowledge the World Climate Research Programme, which, through its Working Group on Coupled Modelling, coordinated and promoted CMIP6. We thank the climate modeling groups for producing and making available their model output, the Earth System Grid Federation (ESGF) for archiving the data and providing access, and the multiple funding agencies who support CMIP6 and ESGF.

## Author Contributions

M.L.C. and E.A.M conceived of the project. M.L.C., K.L., and M.H. performed the data processing and analysis. All authors contributed to analyzing results and writing the paper.

## Data availability

Data is available upon request and code to replicate all results in the main text and supplementary materials is available at https://github.com/marissachilds/global-dengue-temper

## Supplemental Materials

## Supplemental Methods

### Simulated disease dynamics

To understand how this statistical approach will work in a dynamical infectious disease system, we run a toy simulation of disease dynamics with a known temperature response. We then estimate the temperature-dengue relationship using the approach described above. We use a stochastic, compartmental model of transmission that divides the population (N) into susceptible (S), exposed (E), infectious (I), and recovered (R). Model compartments change as follows:

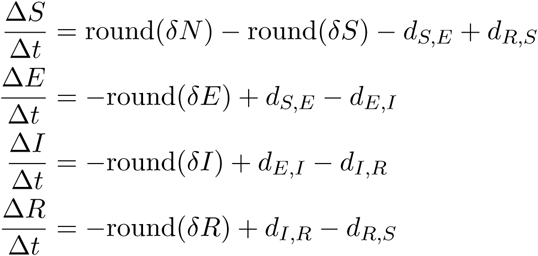

where *δ* is the birth and death rate and *d_X,Y_* is the number of individuals transitioning from class *X* to *Y* . We define the transitions as

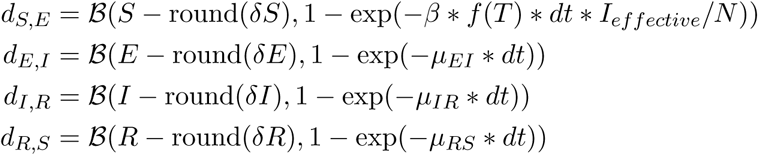

with *B* a simulated binomial process, *µ_XY_* the transition rate between compartments *X* and *Y* , *β* the maximum transmission rate, *f* (*T* ) a function of temperature that ranges from 0 to 1, and *I_effective_* = max(*I,* 1) an effective number of infectious individuals to ensure the pathogen persists in the population. Further, we assume that the total number of reported cases is a a Poisson random variable with mean equal to the total accumulated new infectious individuals over the observation period times some observation rate.

To mimic the nested structure of our data, we run the simulation with 10 countries each with 20 administrative units. We construct temperature data for administrative unit *i* in country *c* on day *d* as a sinusoidal seasonal pattern with daily temperature anomalies

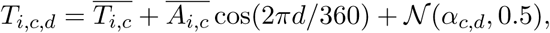

where the unit average temperature 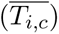 and amplitude 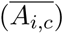 are drawn from distributions centered at the country averages and daily unit temperature anomalies are centered at the daily country temperature anomalies:

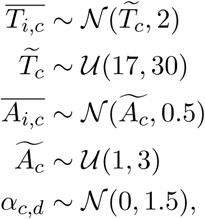

with *U* and *N* uniform and normal distributions, respectively.

For transmission parameters, each country has a transmission rate *β_c_* pulled from a random normal distribution with mean 0.25 and standard deviation 0.04, and each unit within a country has a maximum transmission rate *β_i,c_* pulled from a normal distribution with mean *β_c_* and standard deviation 0.02. We assume the transition rate from exposed to infected (*µ_EI_*) is 1/3 days^−1^, from infected to recovered (*µ_IR_*) is 1/7 days^−1^, and from recovered back to susceptible (*µ_RS_*) is 1/2 years^−1^. The daily birth and death rates (*δ*) are 0.00007 and the population (*N* ) is 1 million individuals. We assume the temperature relationship is *f* (*T* ) = −0.01(*T* − 15)(*T* − 35) for temperatures between 15°C and 35°C and 0 otherwise. To simulate, we initiate the model for each administrative unit with round(U(5, 100)) infected individuals, a portion of the population susceptible—with country level average susceptibility drawn from *S_c,_*_0_ ∼ U(0.5, 0.9) and administrative unit susceptibility drawn from *S_i,c,_*_0_ ∼ N (*S_c,_*_0_, 0.05) and limited to between 0 and 1. The remainder of the population is assumed to be recovered. We fix the country and unit parameters and simulated temperature data as described above, then run the simulations 20 times for each administrative unit. Simulations are run for 20 years using an Euler approximation of the continuous time process using a time step of 1 day using the pomp package in R ^1,2^, and the first 5 years of the simulations are discarded to allow dynamics to stabilize.

We find these simulation produce plausible disease dynamics (Fig. S15) with some locations showing consistent annual outbreaks and others showing larger outbreaks every 2-5 years, consistent with different disease dynamics observed in the dengue data (Fig. 1). First, we use these simulations to estimate a total average temperature response using a similar approach as described above with reported case incidence regressed on cubic polynomials of temperature in the current and previous month. We fit an estimated response for each of the 20 simulations (Fig. S4). Second, we estimate a temperature response interacted with different susceptibility levels, where susceptibility levels are defined using the percent of the population susceptible in the previous month binned into low susceptibility (*<* 60%), medium susceptibility (60-80%) and high susceptibility (*>* 80%) (Fig. S8).

---

**Figure S1:**
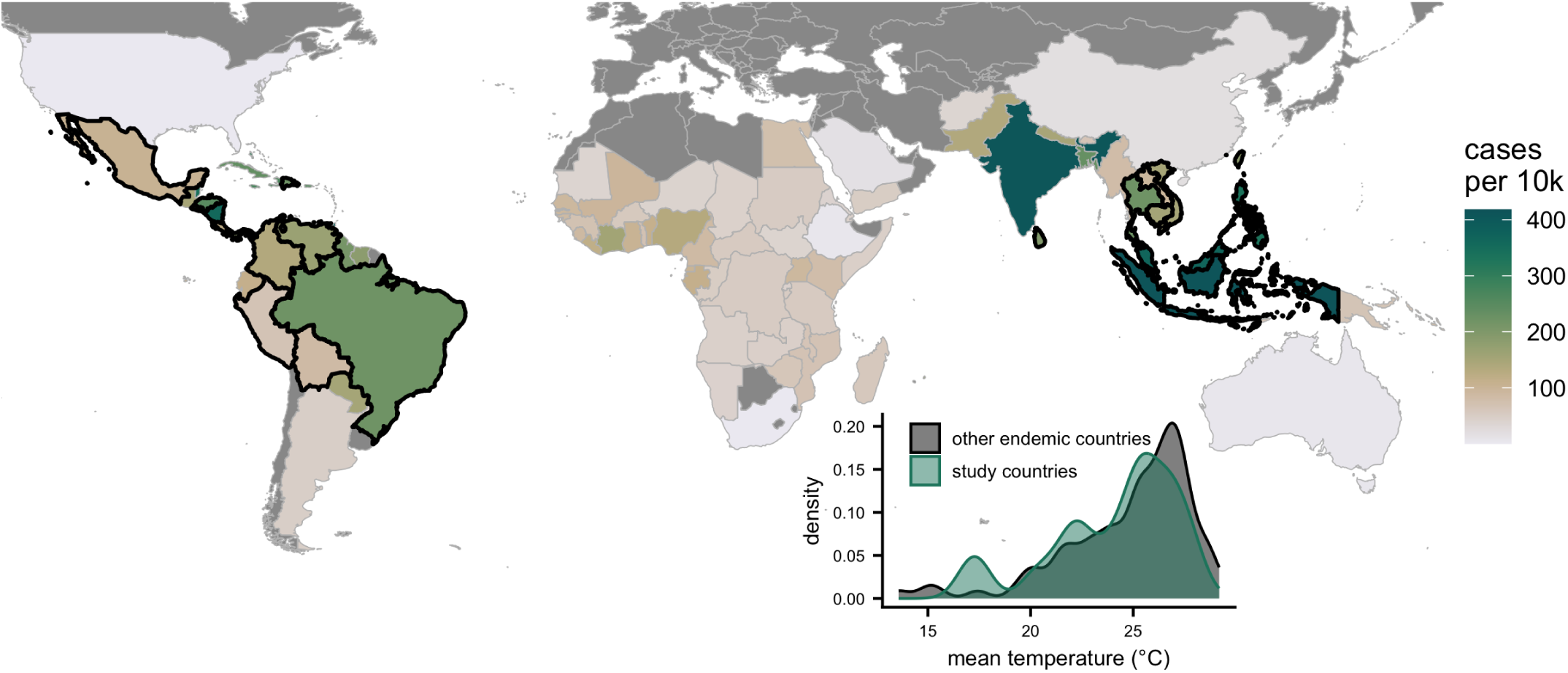
Availability of dengue data in endemic areas. Black borders on the map indicate subnational subannual data was available for that country. Dark grey countries have zero estimated incidence. Dengue cases are annual estimates^3^. Inset shows a density plot of average temperatures in all endemic countries (gray) and in the countries in our dataset (green). We define endemic countries as having greater than 10 cases per 10k based on annual estimated incidence.

**Figure S2:**
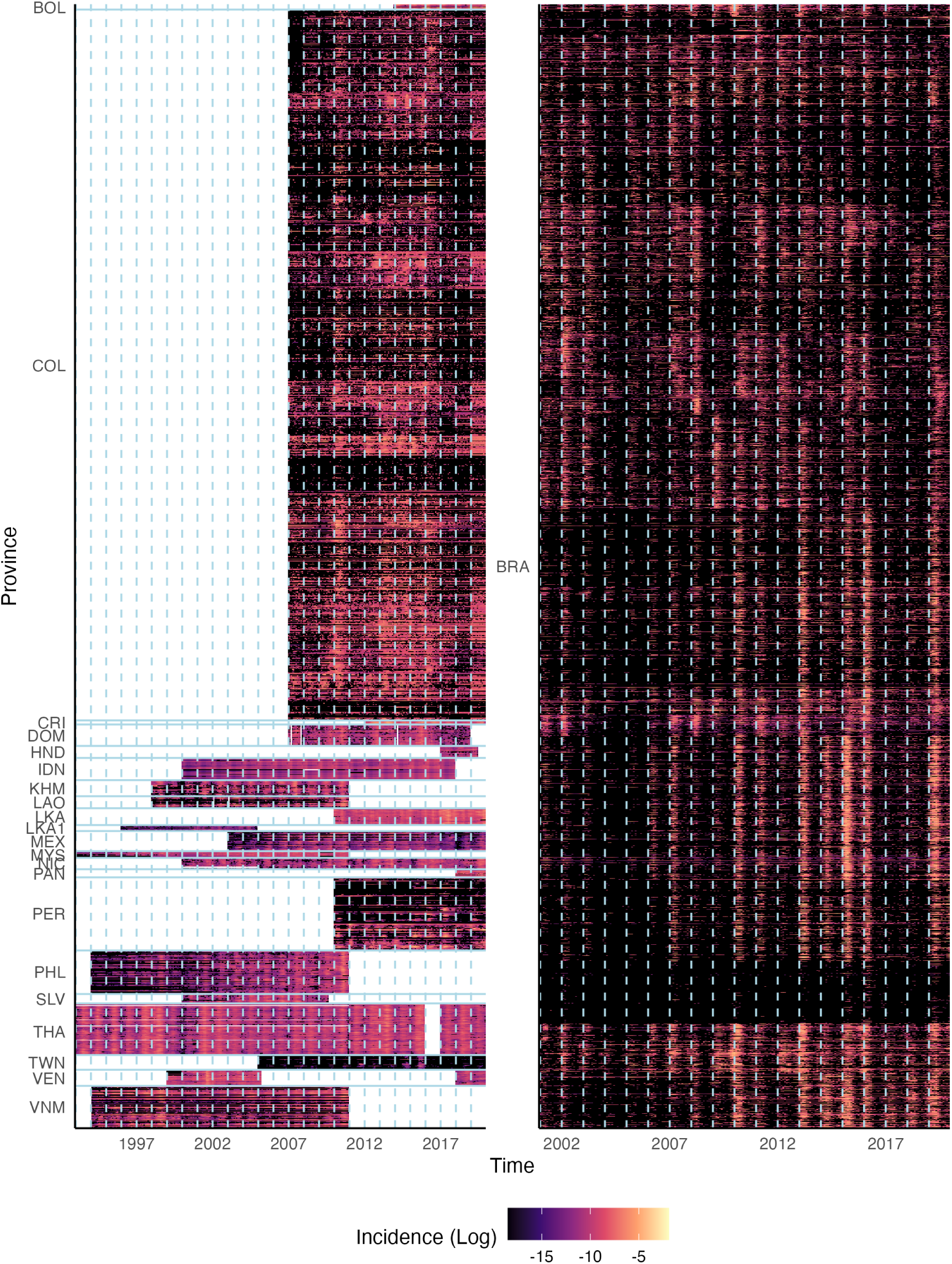
Heatmap of logged monthly dengue incidence and temperature for subnational administrative units. Months with no data are indicated in white while months with no cases are indicated in black. Countries are indicated on the left by their three-letter codes and horizontal lines separate spatial units in different countries. Vertical dashed lines separate years.

**Figure S3:**
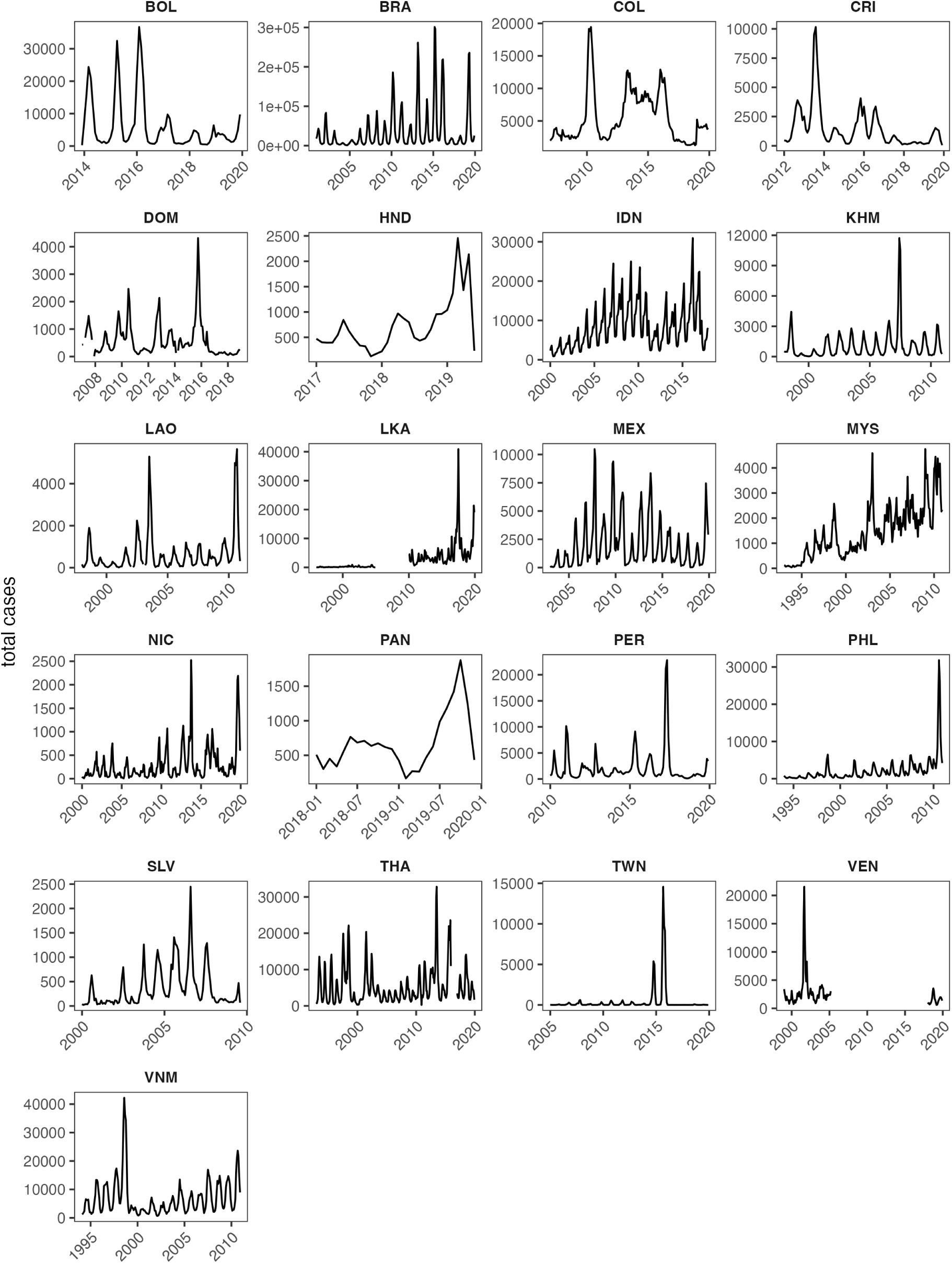
Epidemic dynamics of dengue in each of 21 countries. Monthly time series of dengue incidence by country.

**Figure S4:**
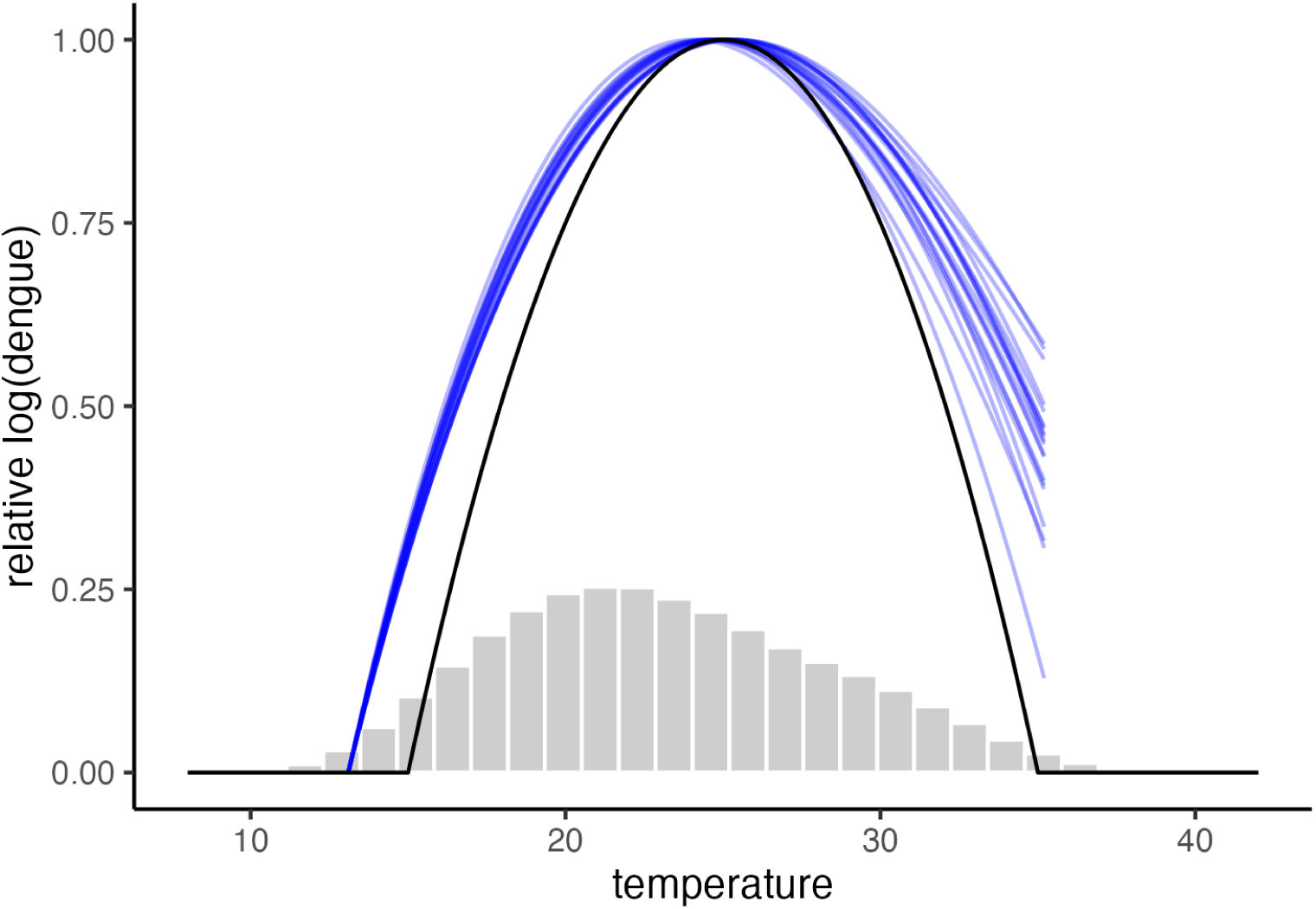
Estimated temperature response from simulated disease dynamics. Black line shows assumed transmission rate - temperature response, the blue lines show the 20 estimated responses scaled between 0 and 1, and the grey histogram shows the distribution of the simulated temperature data. Estimated responses are limited between the 1st and 99th percentiles of the simulated temperature distribution.

**Figure S5:**
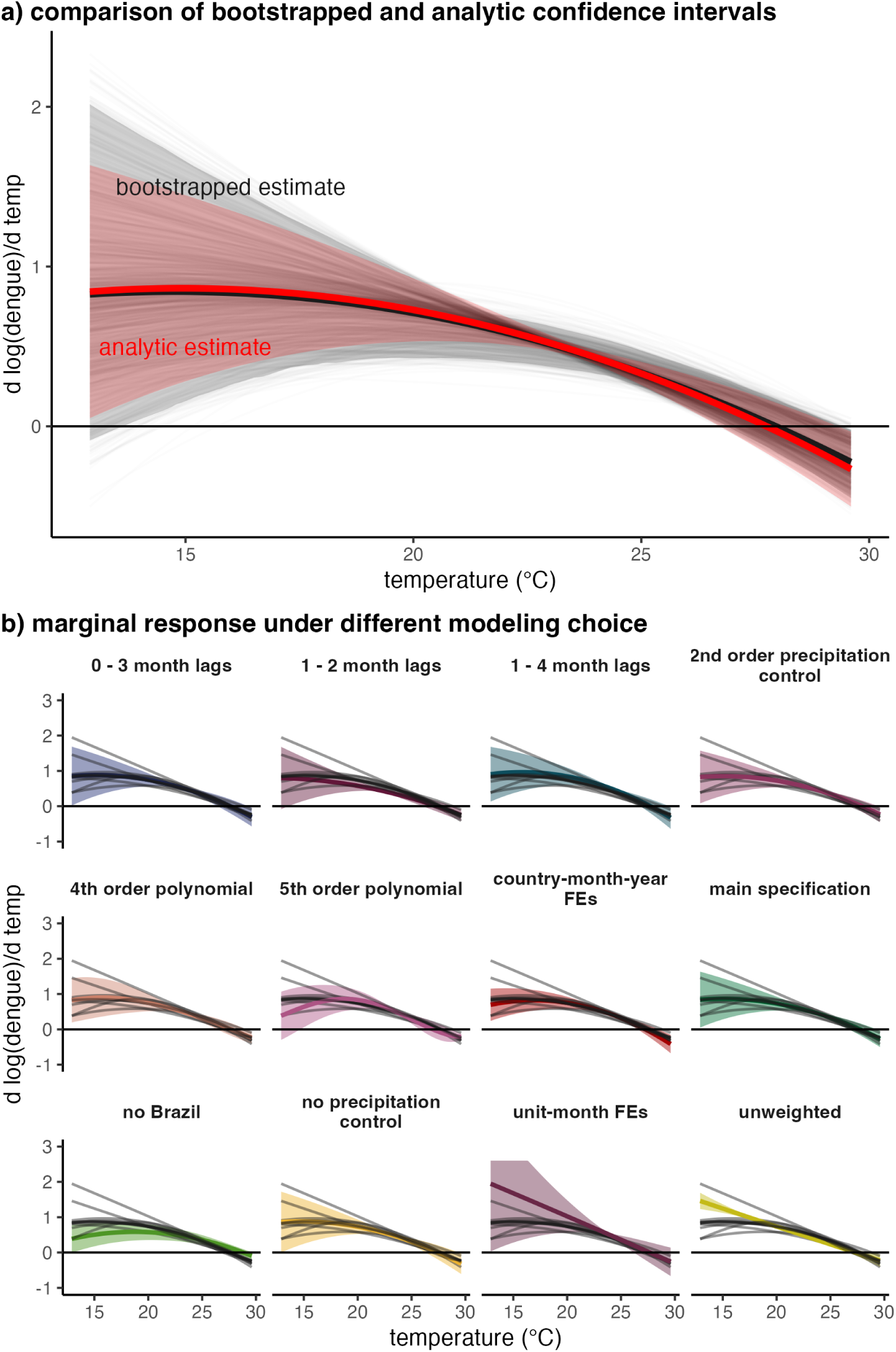
Sensitivity of results to modelling choices. (a) Comparison of 95% confidence intervals produced through bootstrapping (gray shaded region) to those produced through an analytic approach (red shaded region). Individual bootstraps are shown in gray lines. Central estimate for bootstrap is the mean among all bootstraps for each temperature. (b) The marginal response of dengue to temperature under model variations. Except for the bootstrapped estimates noted in panel (a), all central estimates and confidence intervals are from model estimate on the full sample using analytic standard errors clustered at the country.

**Figure S6:**
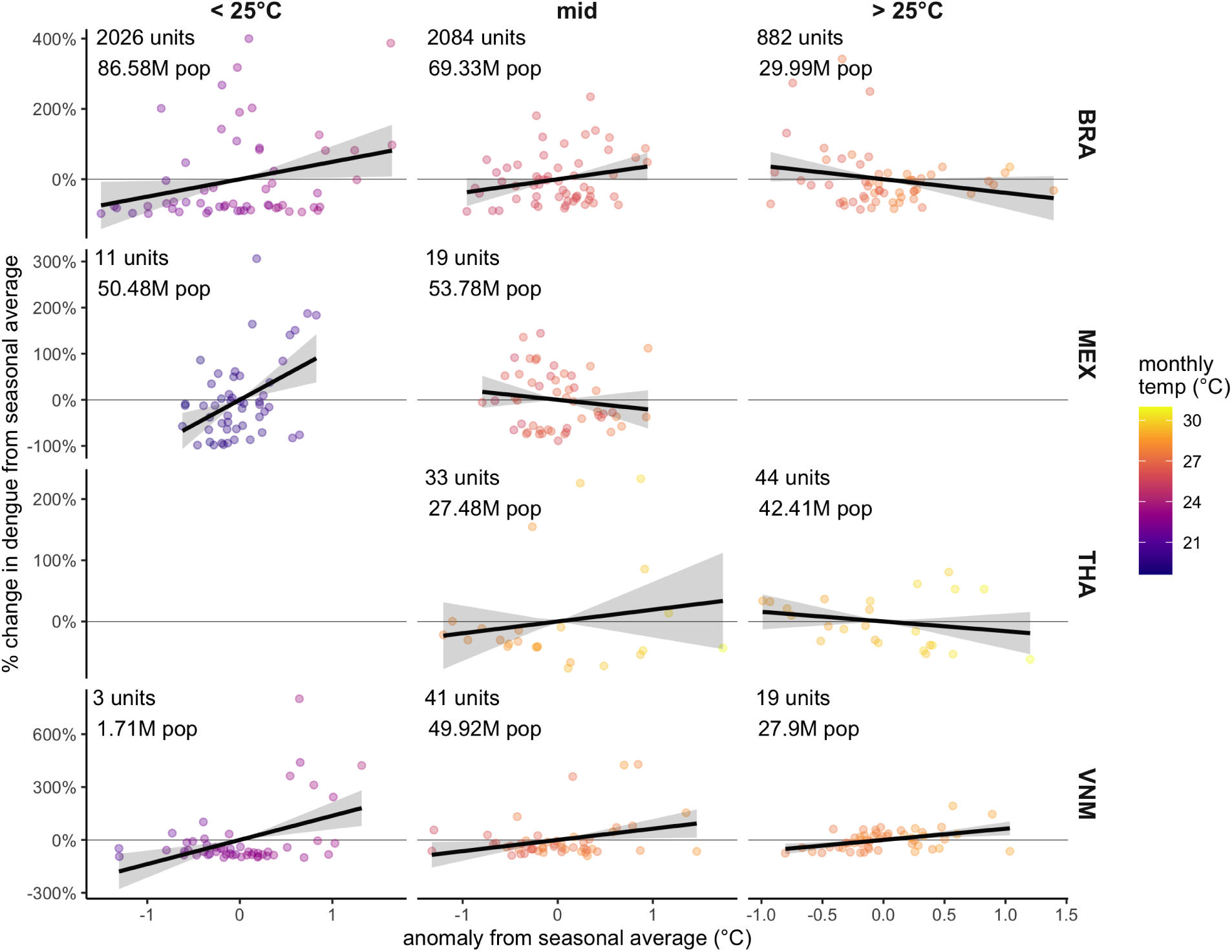
Comparison of seasonal anomalies in dengue and temperature in selected countries. Anomalies are calculated as the absolute (temperature, horizontal axis) or percent change (dengue, vertical axis) from the monthly average in each location during the transmission season. Locations are country aggregates (rows), split into temperature categories (columns) based on season temperatures in sub-country administrative units. Administrative units falling into the *<*25°C category have seasonal average temperatures below 25°C year round, while administrative units in the *>*25°C category have seasonal average temperatures above 25°C year round. The remaining administrative units are grouped into the mid category. Points are colored by the monthly average temperature for each observation. Regression lines shown are from linear models of percent change in dengue anomalies on temperature anomalies without intercept. Transmission season in country-temperature category is defined as the three months per year that have the highest median incidence. Temperature anomalies are 2 months lagged relative to dengue anomalies.

**Figure S7:**
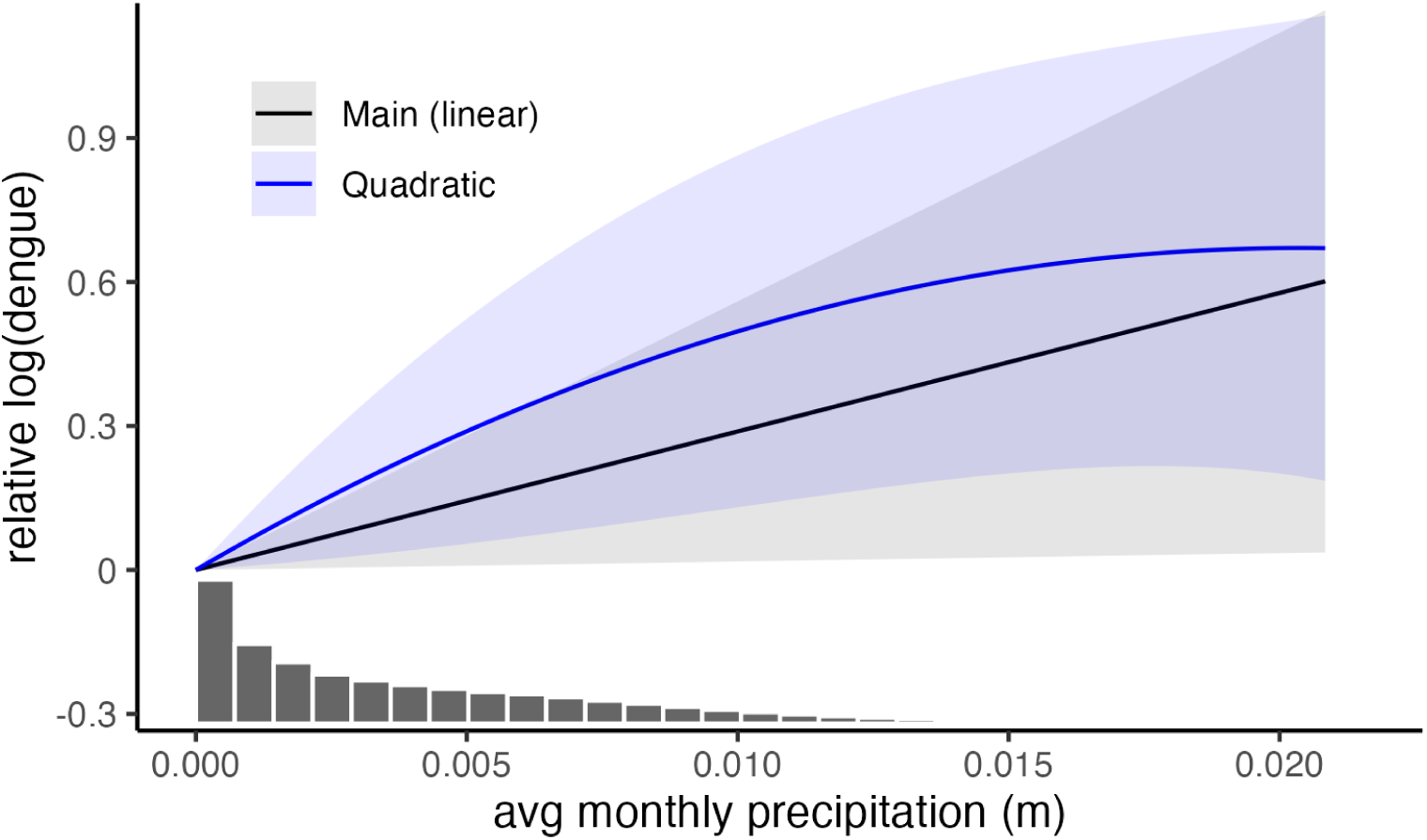
Estimated effect of precipitation on dengue incidence. Estimated effects are from the main model (black) or a model with quadratic precipitation, with central estimates shown by lines and 95% CIs shown by shaded regions calculated from analytic standard errors clustered at the country. Distribution of observed average monthly precipitation values is shown in the grey histogram. Both precipitation responses and distributions are limited to the bottom 99% of the precipitation distribution.

**Figure S8:**
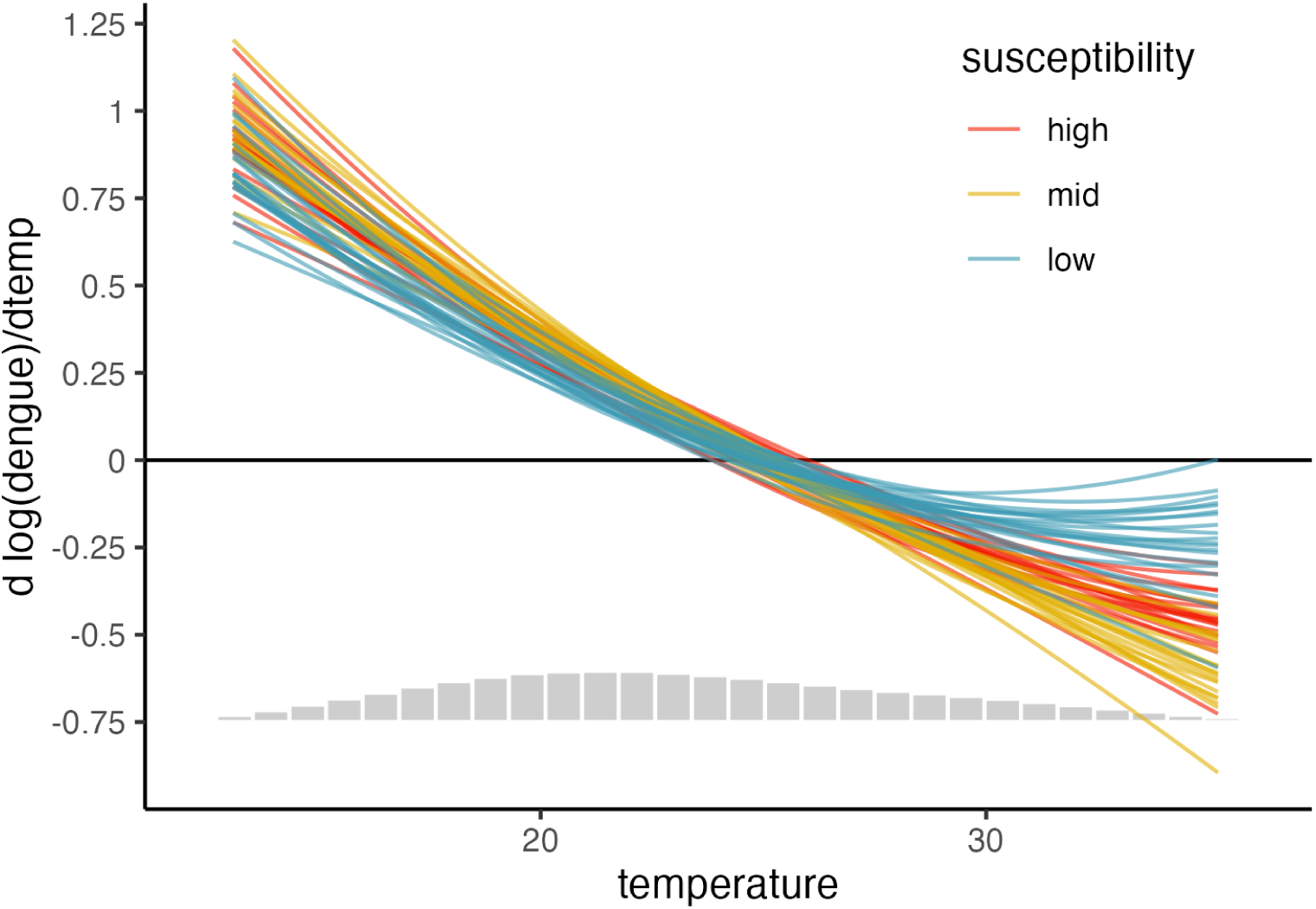
Estimated temperature response from simulated disease dynamics binned by susceptibility levels. The colored lines show the 20 estimated marginal effects for each susceptibility bin (low, *<*60%; mid, 60-80%; high *>*80%) and the grey histogram shows the distribution of the simulated temperature data. Estimated responses are limited between the 1st and 99th percentiles of the simulated temperature distribution.

**Figure S9:**
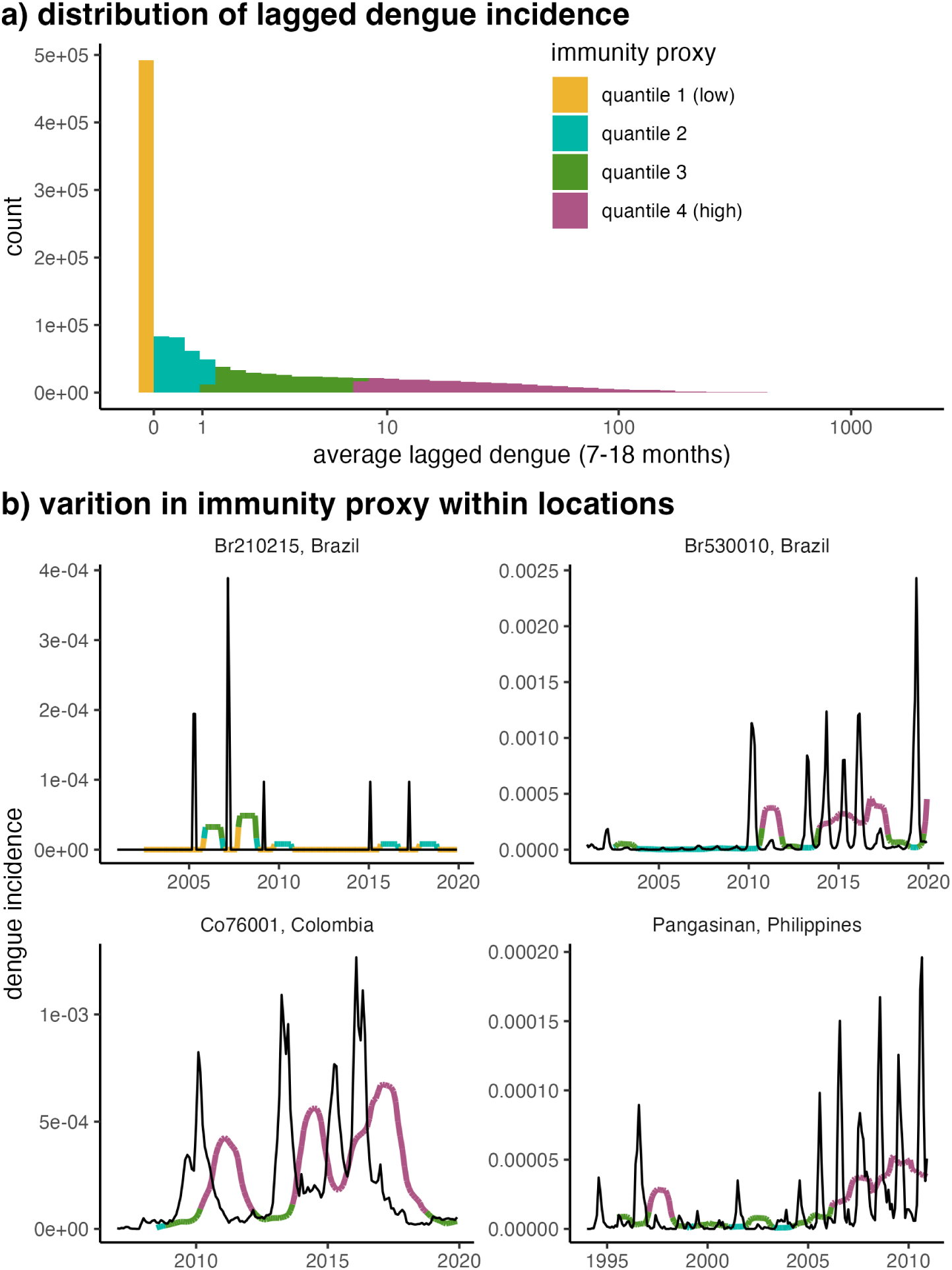
Immunity proxy determined by lagged dengue incidence. a) Quantiles of the immunity proxy are identified from the distribution of average lagged rescaled dengue incidence over the previous 7-18 months. Parts of the distribution are colored by the quantile they fall into. As over more than 25% of the observations had an average lagged dengue incidence = 0, we defined the first quantile as incidence zero, and split the remainder of the distribution into terciles. b) Administrative units show variation in immunity proxy over time, with years with smaller outbreaks more often occurring when the immunity proxy is higher and larger outbreaks more often occurring when the immunity proxy is lower. Black lines show raw dengue incidence for each adminsitrative unit, and colored lines show the average lagged dengue incidence (unscaled to match in scale with observed dengue incidence), with colors indicating the immunity proxy quantile and matching those in a). The four administrative units shown were randomly selected among those with no missing observations, where sampling weights were determined by unit populations.

**Figure S10:**
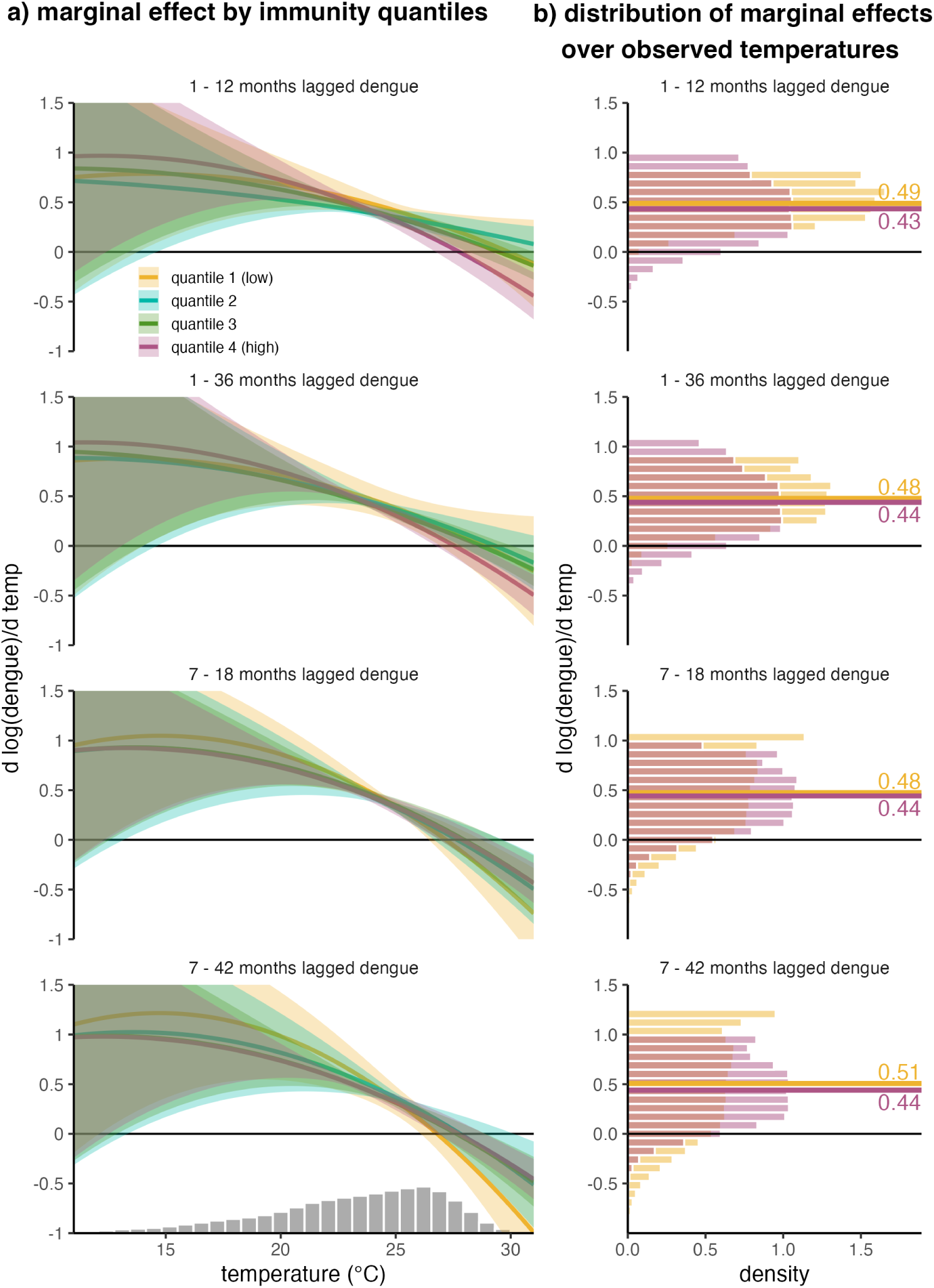
Marginal temperature responses are largely similar across immunity levels. a) Marginal effects of temperature on dengue incidence are estimated for each immunity quantile (color). The grey histrogram shows the distribution of current period (1995 - 2014) temperatures used in climate projections. Lines are central estimates and shaded areas are 95% CIs using analytic standard errors clustered at the country. b) The distribution of marginal effects for the current period temperatures for the lowest (yellow) and highest (purple) immunity quantiles. Histograms show the distribution or marginal effects evaluated at the central temperature response estimate and do not incorporate uncertainty from the marginal response estimates. Horizontal lines mark average estimates for each distribution and text labels indicate the value for each. Each row shows estimates for using average dengue incidence over a different time period as a proxy for immunity.

**Figure S11:**
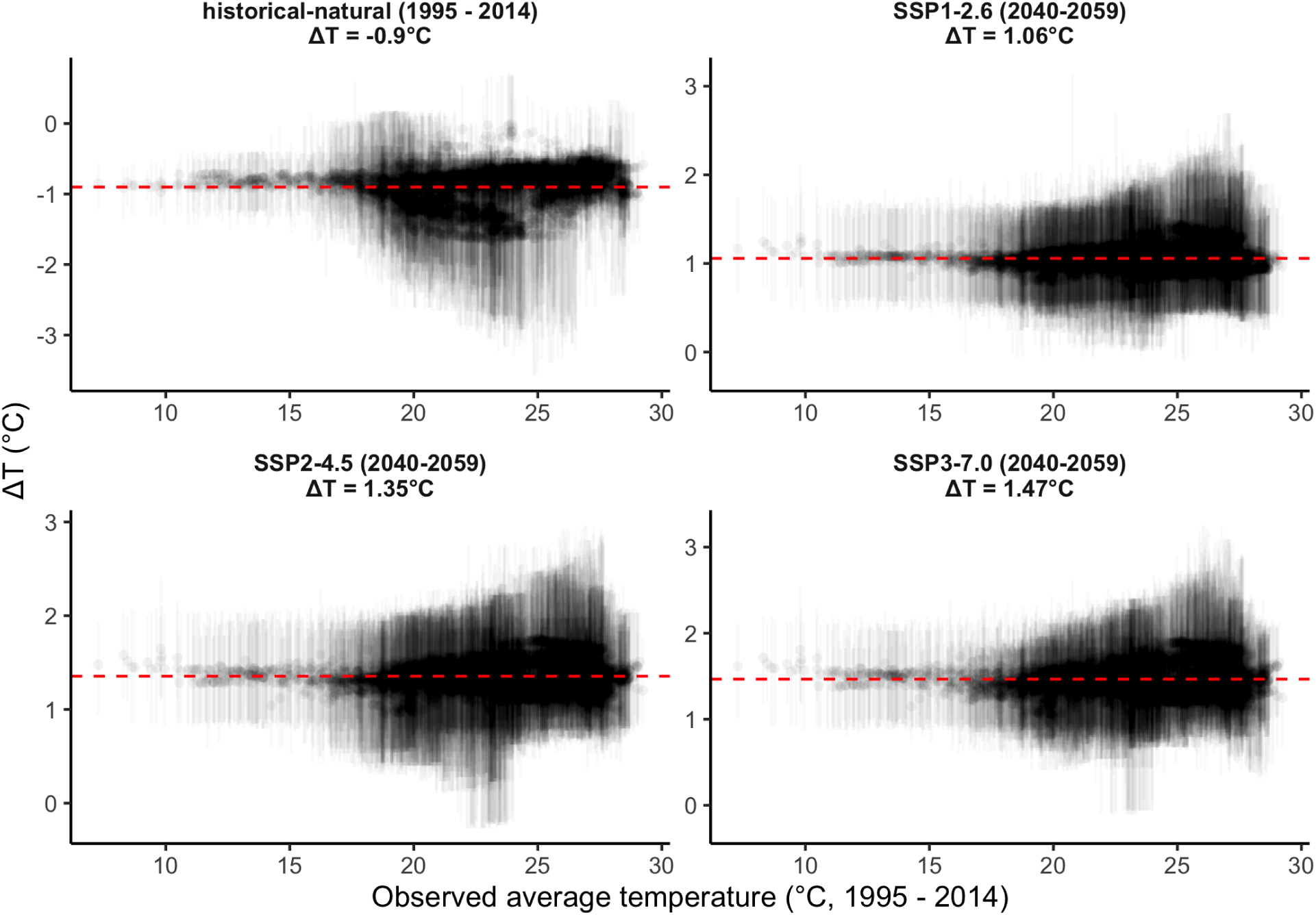
Estimated change in temperature under different climate scenarios. Estimated change in temperature between observed average temperatures (1995 - 2015) and either future climate scenarios (2040 - 2059) or historical climate without anthropogenic forcing (1995 - 2014). Points indicate mean values and line segments indicate 95% CIs over the 21 GCMs. Panel titles indicate the reference time period for the scenario. Average temperature change over all administrative units with reported dengue in the dataset is shown in both panel titles and indicated by the horizontal red line. Note that the scales of the y-axes differ in each panel to span the range of values for that scenario.

**Figure S12:**
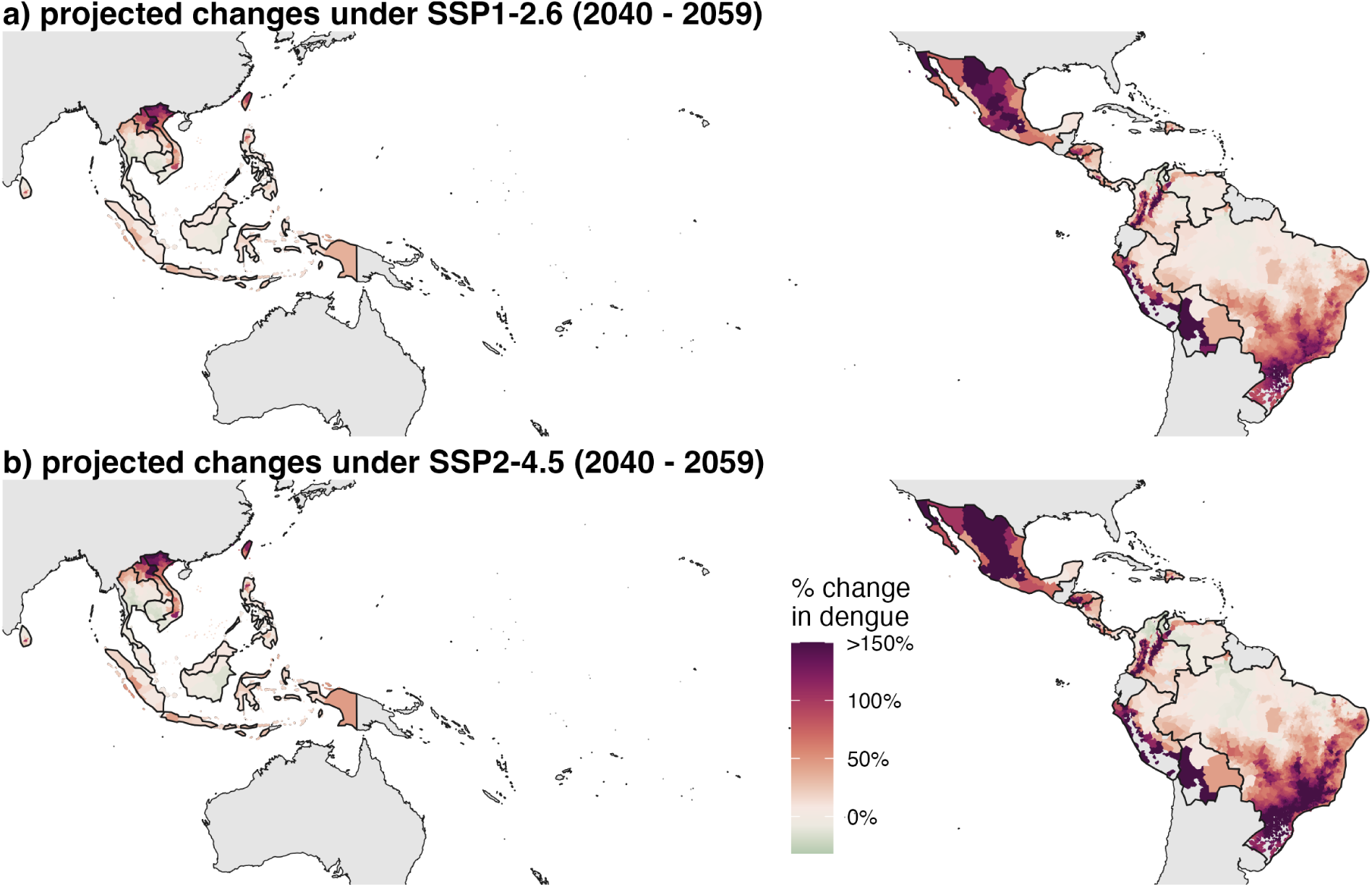
Maps of impacts of future climate warming. Projected change in dengue under climate scenario (1) SPP1-2.6 in 2040-2059 and (b) SPP2-4.5 in 2040-2059 relative to current climate (1995 - 2014).

**Figure S13:**
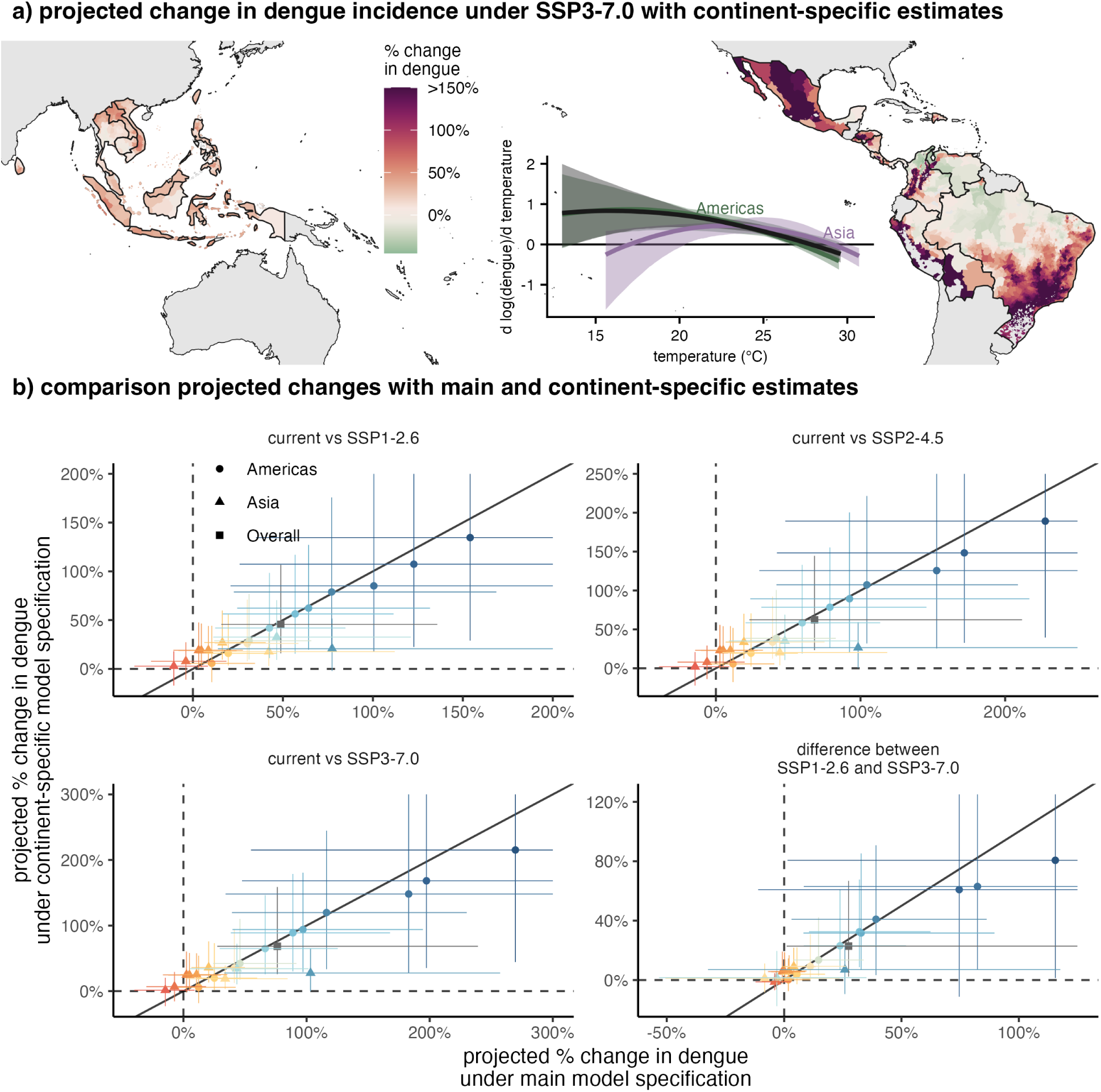
Projections with continent-speficic temperature responses. (a) Projected change in dengue incidence under climate scenario SSP3-7.0 from 2040-2059 based on continentspecific temperature responses. Inset figure shows continent-specific temperature responses for the Americas (green) and Asia (purple) as well as main specification (black) for comparison. (b) Comparison between country average projected changes with the main model (horizontal axis) and continent-specific model (vertical axis) for different future scenarios. Points are mean estimates and line segments indicate 95% CIs. Black diagonal line indicates 1-1. Colors match country colors in Fig. 5. Estimates for Bolivia are ommitted from panel b due to the much larger mean estimated effects and large confidence intervals.

**Figure S14:**
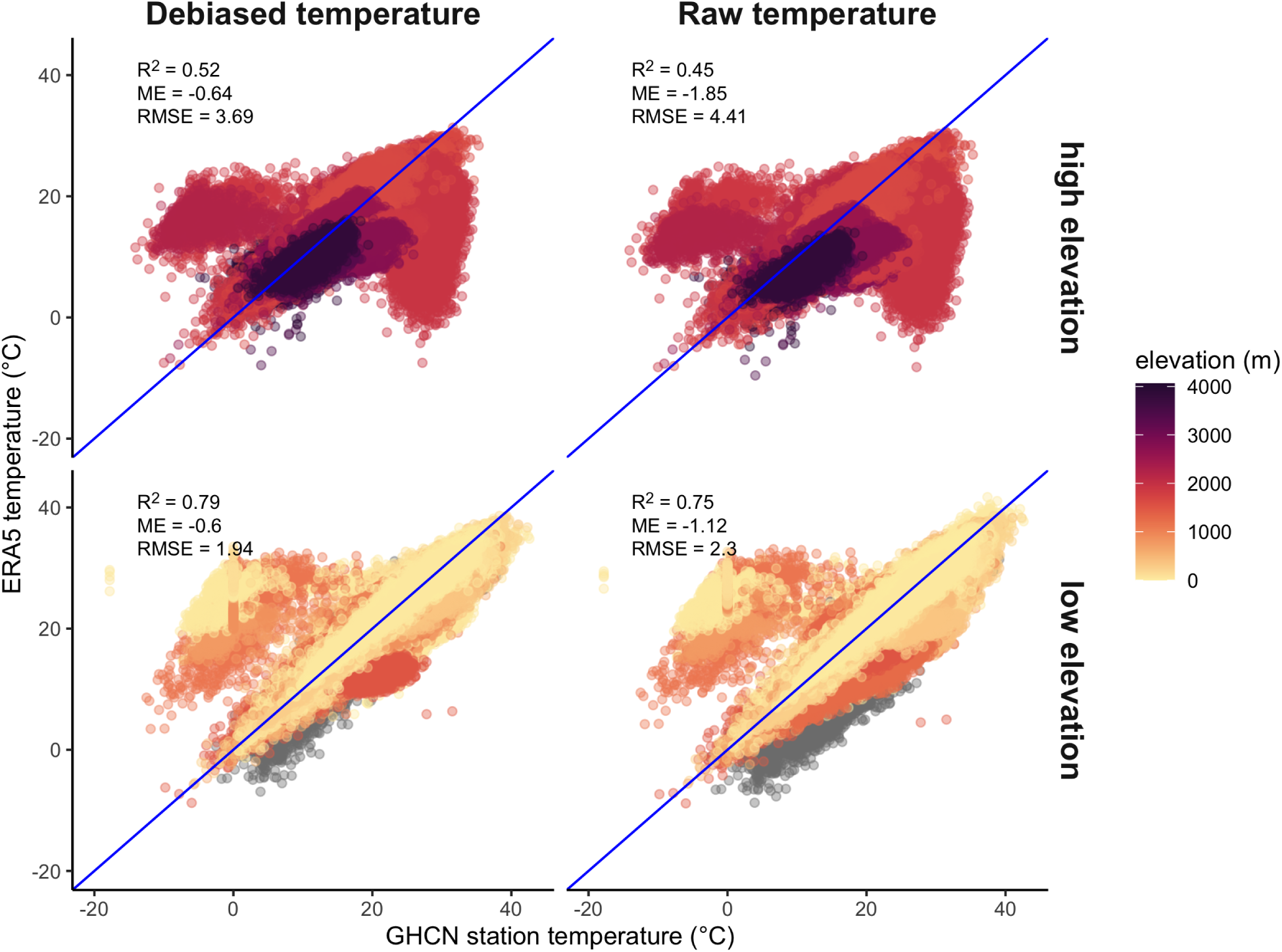
ERA5 temperature is debiased using WorldClim. ERA5 temperature is shown against GHCN weather station data. The ERA5 temperature bias, especially prevalent at high elevations, is reduced using monthly WoldClim climatology. Each point is a station-day with points colored by station elevation. High elevation is defined as stations above 1500 meters. Grey points are missing elevation information in the GHCN data set and are included in the low elevation category. R^2^ from a linear regression of ERA5 temperature on GHCN station temperature, mean error (ME), and root mean squared error (RMSE) are shown in each panel.

**Figure S15:**
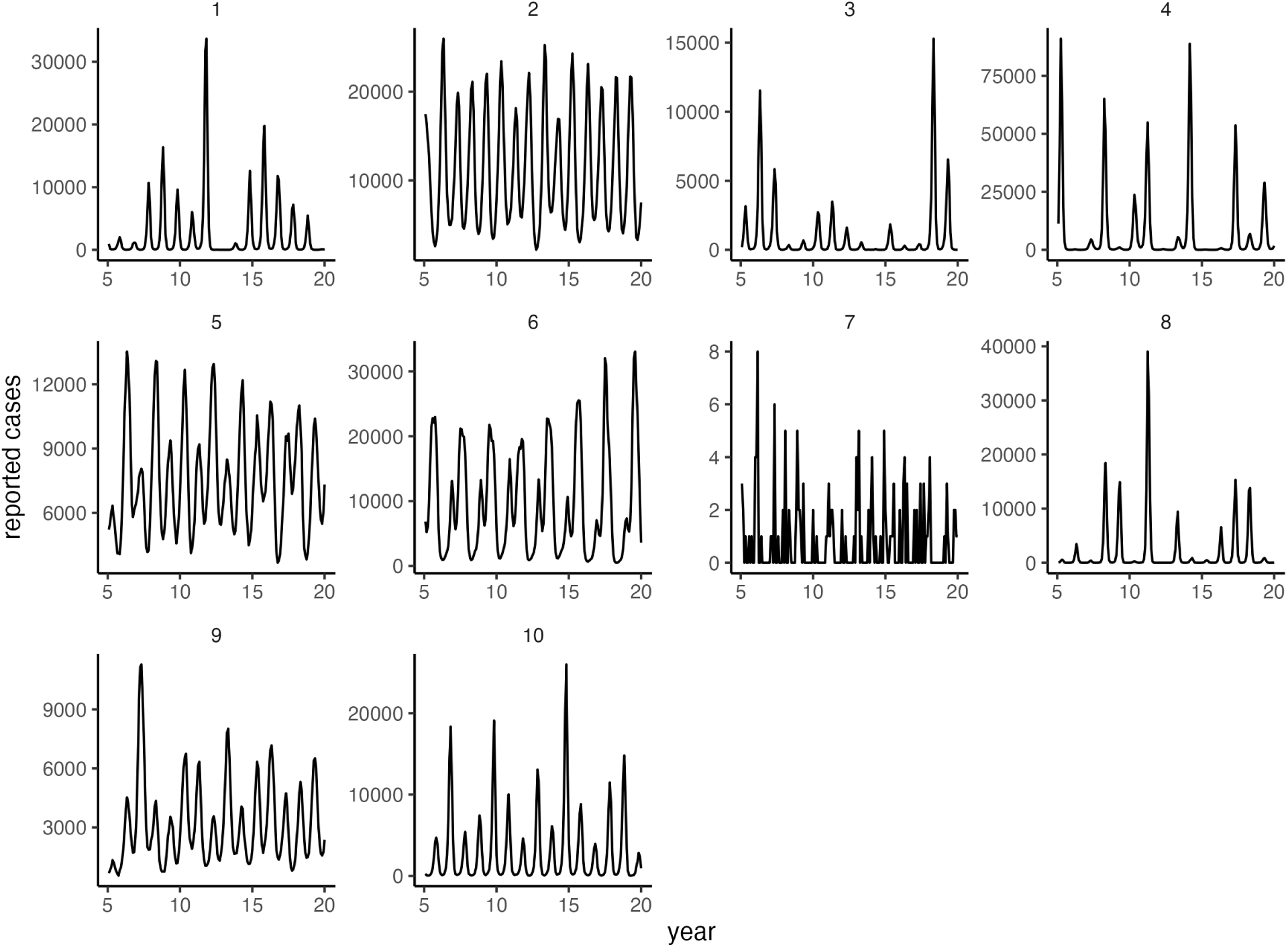
Mechanistic simulations produce plausible disease dynamics. Each panel shows the first simulation for one administrative unit for each simulated county with reported cases on the vertical axis and year on the horizontal axis.

**Table S1:**
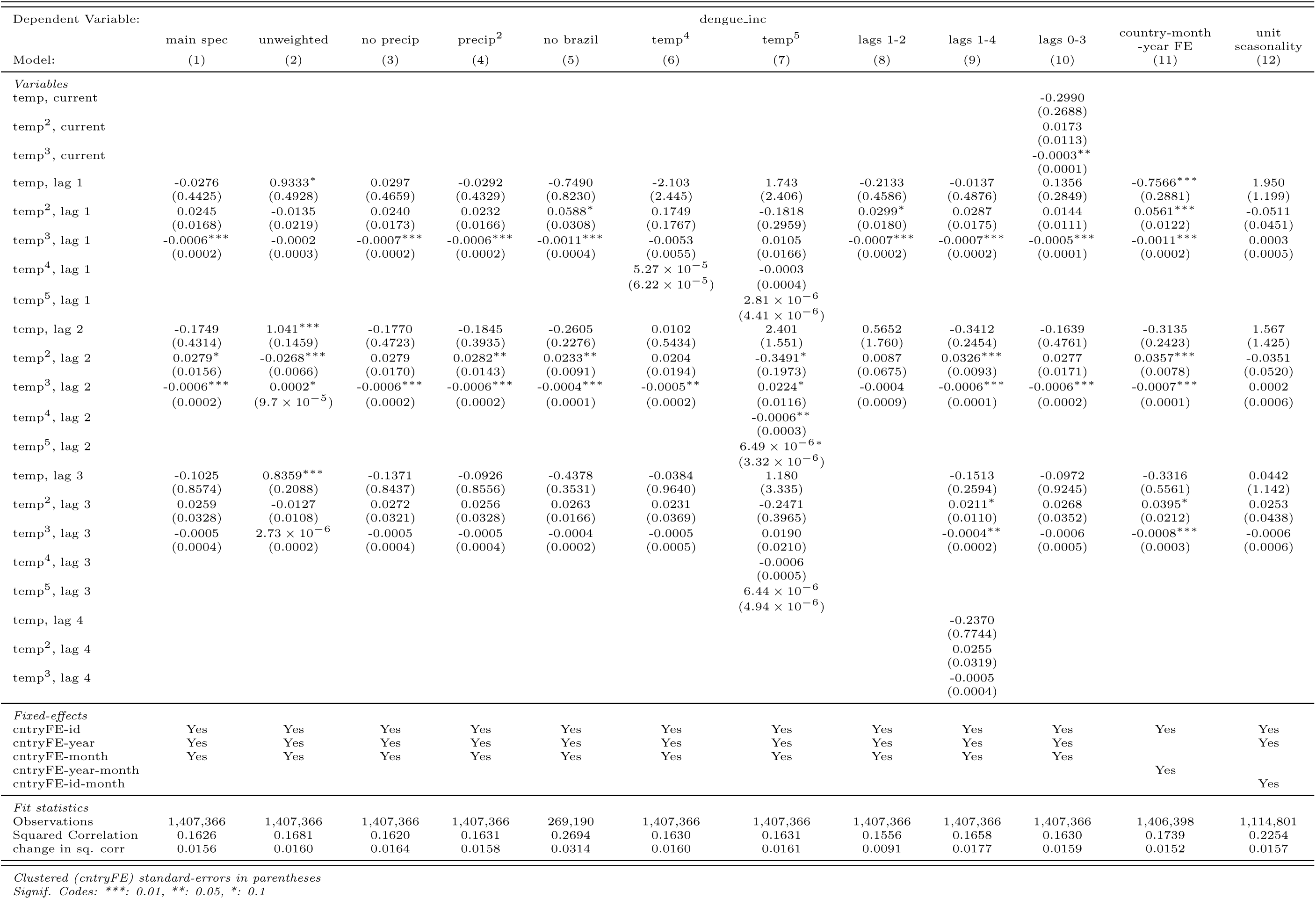
Coefficients from model specifications. Only coefficients for temperature covariates are show. Standard errors are shown in parentheses. Unless noted in the model name, all models use population weights, and linear precipitation controls with the precipitation lags matching the temperature lags. ‘cntryFE’ indicates country-data source fixed effects (see “Estimating dengue-temperature responses”). The “change in sq. corr” statistic measures that change in squared correlation from including the temperature terms.

**Table S2:** Projected percent change in dengue incidence for countries under different climate scenarios. Numbers are mean estimates followed by 95% CIs.

| country | current vs no anthropogenic forcing | current vs SSP1-2.6 | current vs SSP2-4.5 | current vs SSP3-7.0 | difference between SSP1-2.6 and SSP3-7.0 |
| --- | --- | --- | --- | --- | --- |
| BOL | -34.5% (-63.4-16.1%) | 293.6% (12.2-2133.3%) | 563.3% (19-4378%) | 811.1% (22.4-6101.4%) | 517.5% (4-4619%) |
| BRA | -32.6% (-49.8-18.2%) | 64.3% (24.6-131.8%) | 92.4% (23.8-216.7%) | 97.1% (39.8-194.5%) | 32.8% (8.2-89.6%) |
| COL | -29.7% (-45.9-13.2%) | 100.5% (20.9-353.5%) | 152.6% (30-550.9%) | 182.9% (34.2-726%) | 82.4% (8.4-408.6%) |
| CRI | -32.6% (-45.2-20.2%) | 77.1% (22.7-168.6%) | 104.4% (41.9-208.8%) | 116.2% (39.4-230.1%) | 39.1% (3.2-86.3%) |
| DOM | -15.4% (-27.5-6.6%) | 19.6% (7.3-46.6%) | 24.3% (10.2-56.7%) | 25.1% (10.5-59.9%) | 5.5% (-1.1-17.5%) |
| HND | -29.6% (-42.6-17.5%) | 56.8% (16.6-111.5%) | 78.8% (31.4-145.6%) | 88.9% (38.5-167.8%) | 32.1% (10.6-62.4%) |
| IDN | -15.5% (-25.9-8.3%) | 16.3% (6.3-40.5%) | 19.4% (6.8-46.2%) | 20.5% (6.5-49.9%) | 4.1% (-1-13.5%) |
| KHM | 1.2% (-14.2-15.1%) | -10.6% (-32.6-8.3%) | -14.4% (-39.3-9.4%) | -14.7% (-39.9-9.1%) | -4.1% (-12.2-2.6%) |
| LAO | -8.3% (-22.8-3.8%) | 46.5% (11.3-121.1%) | 47.7% (9-99.2%) | 43.4% (5.4-101.6%) | -3.1% (-53.3-35.1%) |
| LKA | -9.9% (-22.1-3.1%) | 8.6% (-1.2-27.8%) | 10% (-1.9-32.6%) | 10.7% (-2.4-35.7%) | 2.1% (-1.7-9.6%) |
| MEX | -27.9% (-46.7-4.6%) | 122.8% (25.9-444.5%) | 171.8% (42.2-571.5%) | 197.4% (47.6-657.9%) | 74.6% (-11.1-304.9%) |
| MYS | -9.1% (-21.5-1.6%) | 3.2% (-10.9-23.9%) | 2.8% (-14.3-27.2%) | 2.4% (-16.7-29.1%) | -0.7% (-6.9-7.7%) |
| NIC | -24.1% (-38.2-14%) | 31.2% (11.8-65.8%) | 41.8% (20-82.9%) | 45.9% (24.1-91.8%) | 14.7% (4-33.9%) |
| PAN | -12.6% (-23.6-6%) | 10.5% (-1.7-34.5%) | 11.9% (-4.3-40.4%) | 12.3% (-4.6-42.4%) | 1.8% (-3.6-11%) |
| PER | -38.7% (-56.3-18.6%) | 154% (35.1-553.4%) | 227.7% (47.9-891%) | 269.6% (54.9-1135.5%) | 115.5% (1.4-581.6%) |
| PHL | -7.6% (-18.3-1.2%) | 4.7% (-8.1-24.3%) | 5.1% (-10.3-28.8%) | 4.9% (-12.3-30.3%) | 0.2% (-5.4-9%) |
| SLV | -35.8% (-53.2-21.3%) | 42.5% (12.3-84.8%) | 59.7% (20.5-113.6%) | 66.3% (29.5-125.4%) | 23.8% (8.3-51.9%) |
| THA | -0.6% (-14.9-9.7%) | -3.9% (-23.3-18.6%) | -6.1% (-26.5-19.9%) | -7.3% (-30.2-18.4%) | -3.3% (-9.6-3%) |
| TWN | -13.2% (-24.1-1.2%) | 77.3% (13.9-208.3%) | 98.3% (25.2-263%) | 103.2% (33.1-257.2%) | 25.9% (-32.4-117.7%) |
| VEN | -18.8% (-29.9-9.8%) | 30.2% (12.1-58.6%) | 39.2% (19.8-75.2%) | 41.4% (21.3-76.5%) | 11.2% (1.5-26.3%) |
| VNM | 0.1% (-13.6-13.3%) | 42.2% (5.8-112.2%) | 44.2% (4.5-118.5%) | 33.9% (2.1-84.6%) | -8.3% (-50.6-23.6%) |
| overall | -18.4% (-27-11.3%) | 48.8% (15.7-135.8%) | 68.2% (23.2-211.7%) | 76.2% (27.4-239.2%) | 27.4% (1.1-125%) |

**Table S3:** Raw dengue data sources and information. MoH = Ministry of Health.

| Country | Spatial resolution | Number of administrative units | Original temporal resolution | Start date | End date | Data source |
| --- | --- | --- | --- | --- | --- | --- |
| BOL | Admin 1 | 9 | weekly | 12/1/13 | 12/31/19 | PAHO |
| BRA | Admin 2 | 5570 | weekly | 1/1/01 | 12/31/19 | MoH DATASUS |
| COL | Admin 2 | 1122 | weekly | 1/1/07 | 12/31/19 | MoH SIVIGILA |
| CRI | Admin 1 | 6 | weekly | 1/1/12 | 12/31/19 | MoH |
| DOM | Admin 2 | 32 | weekly | 1/1/07 | 12/31/18 | MoH DIEPI |
| HND | Admin 1 | 18 | weekly | 1/1/17 | 6/30/19 | PAHO |
| IDN | Admin 1 | 34 | monthly | 1/1/00 | 12/31/17 | Tycho |
| KHM | Admin 1 | 25 | monthly | 1/1/98 | 12/31/10 | Tycho |
| LAO | Admin 1 | 18 | monthly | 1/1/98 | 12/31/10 | Tycho |
| LKA | Admin 2 | 25 | monthly | 1/1/10 | 12/31/19 | MoH Epidemiology Unit |
| LKA | Admin 1 | 9 | monthly | 1/1/96 | 12/31/04 | Tycho |
| MEX | Admin 1 | 32 | weekly | 1/1/03 | 12/31/19 | MoH |
| MYS | Admin 1 | 16 | monthly | 1/1/93 | 12/31/10 | Tycho |
| NIC | Admin 1 | 17 | monthly | 1/1/00 | 12/31/04 | Tycho |
| NIC | Admin 1 | 17 | weekly | 1/1/04 | 12/31/19 | PAHO |
| PAN | Admin 1 | 13 | weekly | 1/1/18 | 12/31/19 | PAHO |
| PER | Admin 2 | 196 | weekly | 1/1/10 | 12/31/19 | CDC Peru |
| PHL | Admin 2 | 87 | monthly | 1/1/94 | 12/31/10 | Tycho |
| SLV | Admin 1 | 14 | monthly | 1/1/00 | 8/31/09 | Tycho |
| THA | Admin 1 | 77 | monthly | 1/1/93 | 12/31/10 | Tycho |
| THA | Admin 1 | 77 | monthly | 1/1/11 | 12/31/19 | MoPH |
| TWN | Admin 2 | 22 | weekly | 1/1/05 | 12/31/19 | Taiwan NIDSS |
| VEN | Admin 1 | 25 | monthly | 1/1/99 | 3/31/05 | Tycho |
| VEN | Admin 1 | 25 | weekly | 1/1/17 | 12/31/19 | PAHO |
| VNM | Admin 1 | 63 | monthly | 1/1/94 | 12/31/10 | Tycho |

**Table S4:**
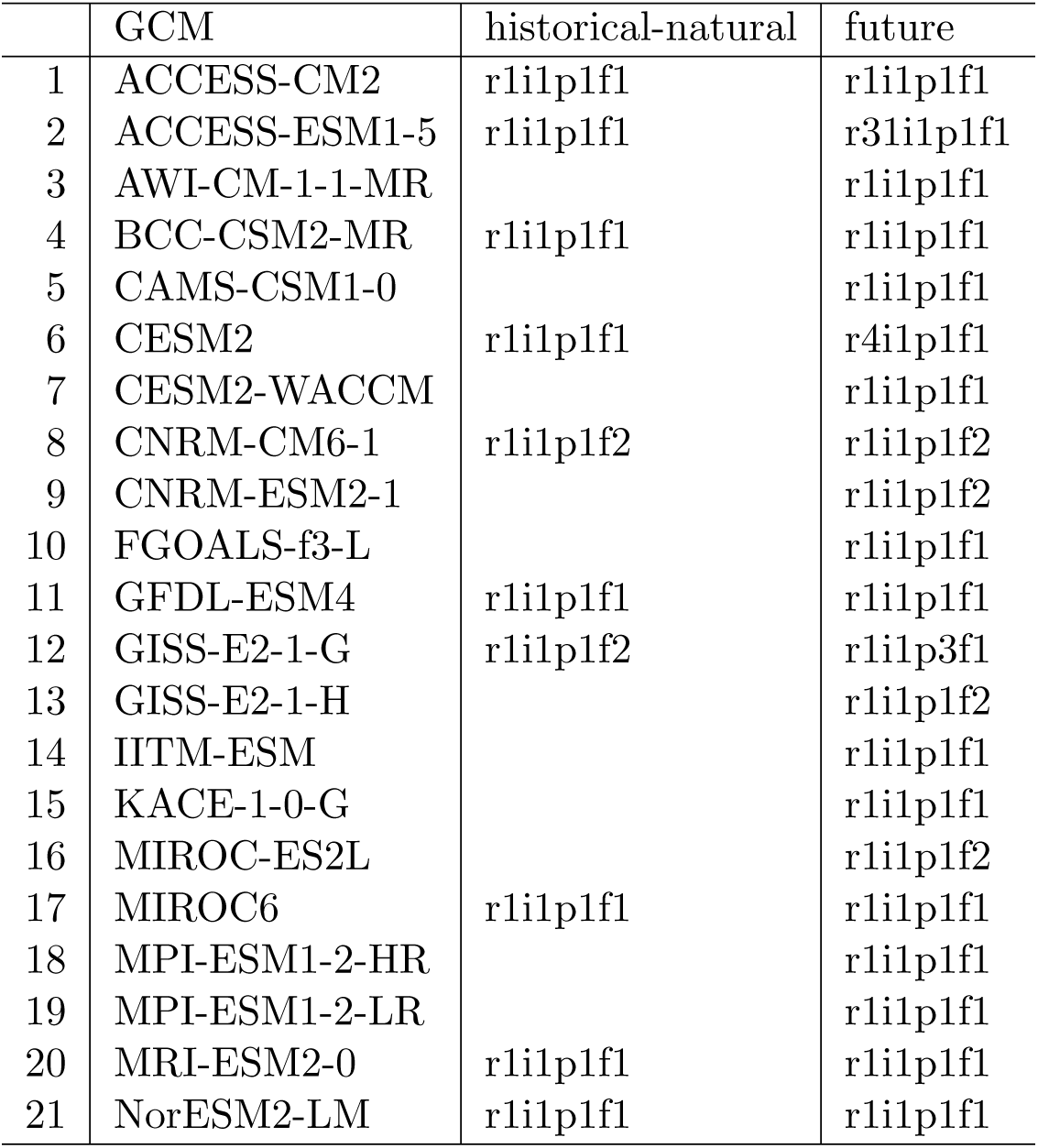
GCM scenarios and variants included in projections. Only a subset of GCMs include a historical-natural scenario.

## References

[1] Kim Knowlton, Miriam Rotkin-Ellman, Linda Geballe, Wendy Max, and Gina M. Solomon. Six Climate Change–Related Events In The United States Accounted For About $14 Billion In Lost Lives And Health Costs. Health Affairs, 30(11):2167–2176, November 2011. ISSN 0278-2715, 1544-5208. doi: 10.1377/hlthaff.2011.0229. URL http://www.healthaffairs.org/doi/10.1377/hlthaff.2011.0229.

[2] Nick Watts, W. Neil Adger, Paolo Agnolucci, Jason Blackstock, Peter Byass, Wenjia Cai, Sarah Chaytor, Tim Colbourn, Mat Collins, Adam Cooper, Peter M. Cox, Joanna Depledge, Paul Drummond, Paul Ekins, Victor Galaz, Delia Grace, Hilary Graham, Michael Grubb, Andy Haines, Ian Hamilton, Alasdair Hunter, Xujia Jiang, Moxuan Li, Ilan Kelman, Lu Liang, Melissa Lott, Robert Lowe, Yong Luo, Georgina Mace, Mark Maslin, Maria Nilsson, Tadj Oreszczyn, Steve Pye, Tara Quinn, My Svensdotter, Sergey Venevsky, Koko Warner, Bing Xu, Jun Yang, Yongyuan Yin, Chaoqing Yu, Qiang Zhang, Peng Gong, Hugh Montgomery, and Anthony Costello. Health and climate change: policy responses to protect public health. The Lancet, 386(10006):1861–1914, November 2015. ISSN 0140-6736, 1474-547X. doi: 10.1016/S0140-6736(15)60854-6. URL https://www.thelancet.com/journals/lancet/article/PIIS0140-6736(15)60854-6/fulltext. Publisher: Elsevier.

[3] Vijay S. Limaye, Wendy Max, Juanita Constible, and Kim Knowlton. Estimating The Costs Of Inaction And The Economic Benefits Of Addressing The Health Harms Of Climate Change: Commentary describes illuminates the costs of inaction on the climate crisis and the economic savings of addressing this problem. Health Affairs, 39(12):2098–2104, December 2020. ISSN 0278-2715, 1544-5208. doi: 10.1377/hlthaff.2020.01109. URL http://www.healthaffairs.org/doi/10.1377/hlthaff.2020.01109.

[4] Jonathan A. Patz, Holly K. Gibbs, Jonathan A. Foley, Jamesine V. Rogers, and Kirk R. Smith. Climate Change and Global Health: Quantifying a Growing Ethical Crisis. EcoHealth, 4(4):397–405, December 2007. ISSN 1612-9202, 1612-9210. doi: 10.1007/s10393-007-0141-1. URL http://link.springer.com/10.1007/s10393-007-0141-1.

[5] Kristie L. Ebi, Christofer Åström, Christopher J. Boyer, Luke J. Harrington, Jeremy J. Hess, Yasushi Honda, Eileen Kazura, Rupert F. Stuart-Smith, and Friederike E. L. Otto. Using Detection And Attribution To Quantify How Climate Change Is Affecting Health. Health Affairs, 39(12):2168–2174, December 2020. ISSN 0278-2715. doi: 10.1377/hlthaff.2020.01004. URL https://www.healthaffairs.org/doi/full/10.1377/hlthaff.2020.01004. Publisher: Health Affairs.

[6] Tamma Carleton, Amir Jina, Michael Delgado, Michael Greenstone, Trevor Houser, Solomon Hsiang, Andrew Hultgren, Robert E Kopp, Kelly E McCusker, Ishan Nath, et al. Valuing the global mortality consequences of climate change accounting for adaptation costs and benefits. The Quarterly Journal of Economics, 137(4):2037–2105, 2022.

[7] Sara Samadzadeh et al. The unfinished agenda of communicable diseases among children and adolescents before the covid-19 pandemic, 1990-2019: A systematic analysis of the global burden of disease study 2019. Lancet, 402(10398):313–335, 2023.

[8] Kevin D Lafferty and Erin A Mordecai. The rise and fall of infectious disease in a warmer world. F1000Research, 5, 2016.

[9] Frank C Tanser, Brian Sharp, and David Le Sueur. Potential effect of climate change on malaria transmission in africa. The lancet, 362(9398):1792–1798, 2003.

[10] Sadie J. Ryan, Colin J. Carlson, Erin A. Mordecai, and Leah R. Johnson. Global expansion and redistribution of Aedes-borne virus transmission risk with climate change. PLOS Neglected Tropical Diseases, 13(3):e0007213, March 2019. ISSN 1935-2735. doi: 10.1371/journal.pntd.0007213. URL https://dx.plos.org/10.1371/journal.pntd.0007213.

[11] Erin A. Mordecai, Jeremy M. Cohen, Michelle V. Evans, Prithvi Gudapati, Leah R. Johnson, Catherine A. Lippi, Kerri Miazgowicz, Courtney C. Murdock, Jason R. Rohr, Sadie J. Ryan, Van Savage, Marta S. Shocket, Anna Stewart Ibarra, Matthew B. Thomas, and Daniel P. Weikel. Detecting the impact of temperature on transmission of Zika, dengue, and chikungunya using mechanistic models. PLOS Neglected Tropical Diseases, 11(4): e0005568, April 2017. ISSN 1935-2735. doi: 10.1371/journal.pntd.0005568. URL https://journals.plos.org/plosntds/article?id=10.1371/journal.pntd.0005568. Publisher: Public Library of Science.

[12] Willem JM Martens, Theo H Jetten, and Dana A Focks. Sensitivity of malaria, schistosomiasis and dengue to global warming. Climatic change, 35(2):145–156, 1997.

[13] Erin A. Mordecai, Jamie M. Caldwell, Marissa K. Grossman, Catherine A. Lippi, Leah R. Johnson, Marco Neira, Jason R. Rohr, Sadie J. Ryan, Van Savage, Marta S. Shocket, Rachel Sippy, Anna M. Stewart Ibarra, Matthew B. Thomas, and Oswaldo Villena. Thermal biology of mosquito-borne disease. Ecology Letters, 22(10):1690–1708, October 2019. ISSN 1461-023X, 1461-0248. doi: 10.1111/ele.13335. URL https://onlinelibrary.wiley.com/doi/10.1111/ele.13335.

[14] Amy Wesolowski, Taimur Qureshi, Maciej F Boni, Pål Roe Sundsøy, Michael A Johansson, Syed Basit Rasheed, Kenth Engø-Monsen, and Caroline O Buckee. Impact of human mobility on the emergence of dengue epidemics in pakistan. Proceedings of the national academy of sciences, 112(38):11887–11892, 2015.

[15] Jane P. Messina, Oliver J. Brady, Nick Golding, Moritz U. G. Kraemer, G. R. William Wint, Sarah E. Ray, David M. Pigott, Freya M. Shearer, Kimberly Johnson, Lucas Earl, Laurie B. Marczak, Shreya Shirude, Nicole Davis Weaver, Marius Gilbert, Raman Velayudhan, Peter Jones, Thomas Jaenisch, Thomas W. Scott, Robert C. Reiner, and Simon I. Hay. The current and future global distribution and population at risk of dengue. Nature Microbiology, 4(9):1508–1515, September 2019. ISSN 2058-5276. doi: 10.1038/s41564-019-0476-8. URL http://www.nature.com/articles/s41564-019-0476-8.

[16] Simon Hales, Neil de Wet, John Maindonald, and Alistair Woodward. Potential effect of population and climate changes on global distribution of dengue fever: an empirical model. The Lancet, 360(9336):830–834, September 2002. ISSN 01406736. doi: 10.1016/S0140-6736(02)09964-6. URL https://linkinghub.elsevier.com/retrieve/pii/S0140673602099646.

[17] Zhiwei Xu, Hilary Bambrick, Francesca D. Frentiu, Gregor Devine, Laith Yakob, Gail Williams, and Wenbiao Hu. Projecting the future of dengue under climate change scenarios: Progress, uncertainties and research needs. PLOS Neglected Tropical Diseases, 14 (3):e0008118, March 2020. ISSN 1935-2735. doi: 10.1371/journal.pntd.0008118. URL https://dx.plos.org/10.1371/journal.pntd.0008118.

[18] Felipe J. Colón-González, Rory Gibb, Kamran Khan, Alexander Watts, Rachel Lowe, and Oliver J. Brady. Projecting the future incidence and burden of dengue in Southeast Asia. Nature Communications, 14(1):5439, September 2023. ISSN 2041-1723. doi: 10.1038/s41467-023-41017-y. URL https://www.nature.com/articles/s41467-023-41017-y.

[19] Colin J. Carlson, Tamma A. Carleton, Romaric C. Odoulami, and Christopher H. Trisos. The historical fingerprint and future impact of climate change on childhood malaria in Africa, July 2023. URL https://www.medrxiv.org/content/10.1101/2023.07.16.23292713v1. Pages: 2023.07.16.23292713.

[20] Jamie M. Caldwell, A. Desiree LaBeaud, Eric F. Lambin, Anna M. Stewart-Ibarra, Bryson A. Ndenga, Francis M. Mutuku, Amy R. Krystosik, Efraín Beltrán Ayala, Assaf Anyamba, Mercy J. Borbor-Cordova, Richard Damoah, Elysse N. Grossi-Soyster, Froilán Heras Heras, Harun N. Ngugi, Sadie J. Ryan, Melisa M. Shah, Rachel Sippy, and Erin A. Mordecai. Climate predicts geographic and temporal variation in mosquito-borne disease dynamics on two continents. Nature Communications, 12(1):1233, February 2021. ISSN 2041-1723. doi: 10.1038/s41467-021-21496-7. URL https://www.nature.com/articles/s41467-021-21496-7.

[21] Nicole Nova, Ethan R. Deyle, Marta S. Shocket, Andrew J. MacDonald, Marissa L. Childs, Martin Rypdal, George Sugihara, and Erin A. Mordecai. Susceptible host availability modulates climate effects on dengue dynamics. Ecology Letters, 24(3):415–425, March 2021. ISSN 1461-023X, 1461-0248. doi: 10.1111/ele.13652. URL https://onlinelibrary.wiley.com/doi/10.1111/ele.13652.

[22] Solomon Hsiang. Climate econometrics. Annual Review of Resource Economics, 8:43–75, 2016.

[23] Zhilin Zeng, Juan Zhan, Liyuan Chen, Huilong Chen, and Sheng Cheng. Global, regional, and national dengue burden from 1990 to 2017: A systematic analysis based on the global burden of disease study 2017. EClinicalMedicine, 32, 2021.

[24] Samir Bhatt, Peter W Gething, Oliver J Brady, Jane P Messina, Andrew W Farlow, Catherine L Moyes, John M Drake, John S Brownstein, Anne G Hoen, Osman Sankoh, et al. The global distribution and burden of dengue. Nature, 496(7446):504–507, 2013.

[25] Rory Gibb, Felipe J. Colón-González, Phan Trong Lan, Phan Thi Huong, Vu Sinh Nam, Vu Trong Duoc, Do Thai Hung, Nguyn Thanh Dong, Vien Chinh Chien, Ly Thi Thuy Trang, Do Kien Quoc, Tran Minh Hoa, Nguyen Hu Tai, Tran Thi Hang, Gina Tsarouchi, Eleanor Ainscoe, Quillon Harpham, Barbara Hofmann, Darren Lumbroso, Oliver J. Brady, and Rachel Lowe. Interactions between climate change, urban infrastructure and mobility are driving dengue emergence in Vietnam. preprint, Epidemiology, July 2023. URL http://medrxiv.org/lookup/doi/10.1101/2023.07.25.23293110.

[26] Devin Kirk, Samantha Straus, Marissa L. Childs, Mallory Harris, Lisa Couper, T. Jonathan Davies, Coreen Forbes, Alyssa-Lois Gehman, Maya L. Groner, Christopher Harley, Kevin D. Lafferty, Van Savage, Eloise Skinner, Mary O’Connor, and Erin A. Mordecai. Temperature impacts on dengue incidence are nonlinear and mediated by climatic and socioeconomic factors. bioRxiv, page 2022.06.15.496305, January 2022. doi: 10.1101/2022.06.15.496305. URL http://biorxiv.org/content/early/2022/06/17/2022.06.15.496305.abstract.

[27] Nicholas G. Reich, Sourya Shrestha, Aaron A. King, Pejman Rohani, Justin Lessler, Siripen Kalayanarooj, In-Kyu Yoon, Robert V. Gibbons, Donald S. Burke, and Derek A. T. Cummings. Interactions between serotypes of dengue highlight epidemiological impact of cross-immunity. Journal of The Royal Society Interface, 10(86):20130414, September 2013. ISSN 1742-5689, 1742-5662. doi: 10.1098/rsif.2013.0414.

[28] Leah C. Katzelnick, Lionel Gresh, M. Elizabeth Halloran, Juan Carlos Mercado, Guillermina Kuan, Aubree Gordon, Angel Balmaseda, and Eva Harris. Antibody-dependent enhancement of severe dengue disease in humans. Science, 358(6365):929–932, November 2017. ISSN 0036-8075, 1095-9203. doi: 10.1126/science.aan6836.

[29] Rosemary A. Aogo, Jose Victor Zambrana, Nery Sanchez, Sergio Ojeda, Guillermina Kuan, Angel Balmaseda, Aubree Gordon, Eva Harris, and Leah C. Katzelnick. Effects of boosting and waning in highly exposed populations on dengue epidemic dynamics. Science Translational Medicine, 15(722):eadi1734, November 2023. ISSN 1946-6234, 1946-6242. doi: 10.1126/scitranslmed.adi1734.

[30] Felipe J. Colón-González, Carlo Fezzi, Iain R. Lake, and Paul R. Hunter. The Effects of Weather and Climate Change on Dengue. PLoS Neglected Tropical Diseases, 7(11):e2503, November 2013. ISSN 1935-2735. doi: 10.1371/journal.pntd.0002503. URL https://dx.plos.org/10.1371/journal.pntd.0002503.

[31] Abdiel E. Laureano-Rosario, Julian E. Garcia-Rejon, Salvador Gomez-Carro, Jose A. Farfan-Ale, and Frank E. Muller-Karger. Modelling dengue fever risk in the State of Yucatan, Mexico using regional-scale satellite-derived sea surface temperature. Acta Tropica, 172: 50–57, August 2017. ISSN 0001706X. doi: 10.1016/j.actatropica.2017.04.017. URL https://linkinghub.elsevier.com/retrieve/pii/S0001706X1730089X.

[32] Christine Giesen, Jesús Roche, Lidia Redondo-Bravo, Claudia Ruiz-Huerta, Diana Gomez- Barroso, Agustin Benito, and Zaida Herrador. The impact of climate change on mosquitoborne diseases in Africa. Pathogens and Global Health, 114(6):287–301, August 2020. ISSN 2047-7724, 2047-7732. doi: 10.1080/20477724.2020.1783865. URL https://www.tandfonline.com/doi/full/10.1080/20477724.2020.1783865.

[33] Erin A Mordecai, Sadie J Ryan, Jamie M Caldwell, Melisa M Shah, and A Desiree LaBeaud. Climate change could shift disease burden from malaria to arboviruses in africa. The Lancet Planetary Health, 4(9):e416–e423, 2020.

[34] Kevin Louis Bardosh, Sadie J. Ryan, Kris Ebi, Susan Welburn, and Burton Singer. Addressing vulnerability, building resilience: community-based adaptation to vector-borne diseases in the context of global change. Infectious Diseases of Poverty, 6(1):166, December 2017. ISSN 2049-9957. doi: 10.1186/s40249-017-0375-2. URL 10.1186/s40249-017-0375-2.

[35] Oliver J. Brady and Simon I. Hay. The Global Expansion of Dengue: How *Aedes aegypti* Mosquitoes Enabled the First Pandemic Arbovirus. Annual Review of Entomology, 65(1):191–208, January 2020. ISSN 0066-4170, 1545-4487. doi: 10.1146/annurev-ento-011019-024918. URL https://www.annualreviews.org/doi/10.1146/annurev-ento-011019-024918.

[36] Aidsa Rivera, Laura E. Adams, Tyler M. Sharp, Jennifer A. Lehman, Stephen H. Waterman, and Gabriela Paz-Bailey. Travel-Associated and Locally Acquired Dengue Cases — United States, 2010–2017. Morbidity and Mortality Weekly Report, 69(6):149–154, February 2020. ISSN 0149-2195. doi: 10.15585/mmwr.mm6906a1. URL https://www.ncbi.nlm.nih.gov/pmc/articles/PMC7017959/.

[37] Céline M Gossner, Nelly Fournet, Christina Frank, Beatriz Fernández-Martínez, Martina Del Manso, Joana Gomes Dias, and Henriette de Valk. Dengue virus infections among european travellers, 2015 to 2019. Eurosurveillance, 27(2):2001937, 2022.

[38] Tilly Alcayna, Isabel Fletcher, Rory Gibb, Léo Tremblay, Sebastian Funk, Bhargavi Rao, and Rachel Lowe. Climate-sensitive disease outbreaks in the aftermath of extreme climatic events: A scoping review. One Earth, 5(4):336–350, April 2022. ISSN 2590-3322. doi: 10.1016/j.oneear.2022.03.011. URL https://www.sciencedirect.com/science/article/pii/S2590332222001440.

[39] Mallory J Harris, Kevin S Martel, César V Munyaco, Andrés G Lescano, Erin A Mordecai, Jared T Trok, Noah S Diffenbaugh, and Mercy J Borbor Cordova. Extreme precipitation, exacerbated by anthropogenic climate change, drove peru’s record-breaking 2023 dengue out-break. medRxiv, pages 2024–10, 2024.

[40] MS Mustafa, V Rasotgi, S Jain, and VJMJAFI Gupta. Discovery of fifth serotype of dengue virus (denv-5): A new public health dilemma in dengue control. Medical journal armed forces India, 71(1):67–70, 2015.

[41] Lise Alves. Brazil to start widespread dengue vaccinations. The Lancet, 403(10422):133, January 2024. ISSN 0140-6736. doi: 10.1016/S0140-6736(24)00046-1. URL https://www.sciencedirect.com/science/article/pii/S0140673624000461.

[42] Hervé Douville, Krishnan Raghavan, James Renwick, Richard P Allan, Paola A Arias, Mathew Barlow, Ruth Cerezo-Mota, Annalisa Cherchi, ThianY Gan, Joëlle Gergis, et al. Water cycle changes. 2021.

[43] Nicholas H. Ogden. Climate change and vector-borne diseases of public health significance. FEMS Microbiology Letters, 364(19):fnx186, October 2017. ISSN 0378-1097. doi: 10.1093/femsle/fnx186. URL 10.1093/femsle/fnx186.

[44] Jan C. Semenza and Shlomit Paz. Climate change and infectious disease in Europe: Impact, projection and adaptation. The Lancet Regional Health - Europe, 9:100230, October 2021. ISSN 26667762. doi: 10.1016/j.lanepe.2021.100230. URL https://linkinghub.elsevier.com/retrieve/pii/S2666776221002167.

[45] ESRI ArcGIS Hub. World cities. Accessed from https://hub.arcgis.com/datasets/esri::world-cities/about on September 26, 2023.

## References

[1] Robin Edwards, Maksym Bondarenko, Andrew Tatem, and Alessandro Sorichetta. Unconstrained national Population Weighted Density in 2000, 2005, 2010, 2015 and 2020 ( 1km resolution ), 2021. URL https://www.worldpop.org/doi/10.5258/SOTON/WP00702.

[2] Noel Gorelick, Matt Hancher, Mike Dixon, Simon Ilyushchenko, David Thau, and Rebecca Moore. Google earth engine: Planetary-scale geospatial analysis for everyone. Remote Sensing of Environment, 2017. doi: 10.1016/j.rse.2017.06.031. URL 10.1016/j.rse.2017.06.031.

[3] United Nations Office for the Coordination of Humanitarian Affairs. Humanitarian Data Exchange. https://data.humdata.org/.

[4] Robert J. Hijmans. Second-level administrative divisions, taiwan, 2015. [shapefile]. University of California, Berkeley. Museum of Vertebrate Zoology, 2015. Retrieved from https://earthworks.stanford.edu/catalog/stanford-fn648mm8787.

[5] Socio-economic regions of Costa Rica — second.wiki. https://second.wiki/wiki/regiones_socioeconc3b3micas_de_costa_rica. [Accessed 08-01-2024].

[6] Copernicus Climate Change Service (C3S) (2017). Era5: Fifth generation of ecmwf atmospheric reanalyses of the global climate. Copernicus Climate Change Service Climate Data Store (CDS), https://cds.climate.copernicus.eu/cdsapp#!/home.

[7] Stephen E. Fick and Robert J. Hijmans. WorldClim 2: new 1-km spatial resolution climate surfaces for global land areas. International Journal of Climatology, 37(12):4302–4315, October 2017. ISSN 0899-8418, 1097-0088. doi: 10.1002/joc.5086. URL https://onlinelibrary.wiley.com/doi/10.1002/joc.5086.

[8] Coupled model intercomparison project 6. Accessed from https://registry.opendata.aws/cmip6.

[9] Veronika Eyring, Sandrine Bony, Gerald A Meehl, Catherine A Senior, Bjorn Stevens, Ronald J Stouffer, and Karl E Taylor. Overview of the coupled model intercomparison project phase 6 (cmip6) experimental design and organization. Geoscientific Model Development, 9(5):1937–1958, 2016.

[10] Intergovernmental Panel On Climate Change. Climate Change 2021 – The Physical Science Basis: Working Group I Contribution to the Sixth Assessment Report of the Intergovernmental Panel on Climate Change. Cambridge University Press, 1 edition, July 2023. ISBN 978-1-00-915789-6. doi: 10.1017/9781009157896. URL https://www.cambridge.org/core/product/identifier/9781009157896/type/book.

[11] Zeke Hausfather, Kate Marvel, Gavin A. Schmidt, John W. Nielsen-Gammon, and Mark Zelinka. Climate simulations: recognize the ‘hot model’ problem. Nature, 605(7908): 26–29, May 2022. ISSN 0028-0836, 1476-4687. doi: 10.1038/d41586-022-01192-2. URL https://www.nature.com/articles/d41586-022-01192-2.

[12] Gerald A. Meehl, Catherine A. Senior, Veronika Eyring, Gregory Flato, Jean-Francois Lamarque, Ronald J. Stouffer, Karl E. Taylor, and Manuel Schlund. Context for interpreting equilibrium climate sensitivity and transient climate response from the CMIP6 Earth system models. Science Advances, 6(26):eaba1981, June 2020. ISSN 2375-2548. doi: 10.1126/sciadv.aba1981. URL https://www.science.org/doi/10.1126/sciadv.aba1981.

[13] Giovanni Di Virgilio, Fei Ji, Eugene Tam, Nidhi Nishant, Jason P. Evans, Chris Thomas, Matthew L. Riley, Kathleen Beyer, Michael R. Grose, Sugata Narsey, and Francois Delage. Selecting CMIP6 GCMs for CORDEX Dynamical Downscaling: Model Performance, Independence, and Climate Change Signals. Earth’s Future, 10(4):e2021EF002625, April 2022. ISSN 2328-4277, 2328-4277. doi: 10.1029/2021EF002625. URL https://agupubs.onlinelibrary.wiley.com/doi/10.1029/2021EF002625.

[14] Alejandro Di Luca, Andrew J. Pitman, and Ramón de Elía. Decomposing Temperature Extremes Errors in CMIP5 and CMIP6 Models. Geophysical Research Letters, 47(14): e2020GL088031, July 2020. ISSN 0094-8276, 1944-8007. doi: 10.1029/2020GL088031. URL https://agupubs.onlinelibrary.wiley.com/doi/10.1029/2020GL088031.

[15] David S. Schoeman, Alex Sen Gupta, Cheryl S. Harrison, Jason D. Everett, Isaac Brito- Morales, Lee Hannah, Laurent Bopp, Patrick R. Roehrdanz, and Anthony J. Richardson. Demystifying global climate models for use in the life sciences. Trends in Ecology & Evolution, 38(9):843–858, September 2023. ISSN 01695347. doi: 10.1016/j.tree.2023.04.005. URL https://linkinghub.elsevier.com/retrieve/pii/S016953472300085X.

[16] World Health Organization Global Health Expenditure database (apps.who.int/nha/database). Accessed on June 14, 2022 from https://data.worldbank.org/indicator/SH.XPD.EHEX.PP.CD.

[17] Zhilin Zeng, Juan Zhan, Liyuan Chen, Huilong Chen, and Sheng Cheng. Global, regional, and national dengue burden from 1990 to 2017: A systematic analysis based on the global burden of disease study 2017. EClinicalMedicine, 32, 2021.

[18] Laurent Bergé. Efficient estimation of maximum likelihood models with multiple fixedeffects: the R package FENmlm. CREA Discussion Papers, (13), 2018.

[19] Nicholas G. Reich, Sourya Shrestha, Aaron A. King, Pejman Rohani, Justin Lessler, Siripen Kalayanarooj, In-Kyu Yoon, Robert V. Gibbons, Donald S. Burke, and Derek A. T. Cummings. Interactions between serotypes of dengue highlight epidemiological impact of cross-immunity. Journal of The Royal Society Interface, 10(86):20130414, September 2013. ISSN 1742-5689, 1742-5662. doi: 10.1098/rsif.2013.0414.

[20] Leah C. Katzelnick, Lionel Gresh, M. Elizabeth Halloran, Juan Carlos Mercado, Guillermina Kuan, Aubree Gordon, Angel Balmaseda, and Eva Harris. Antibody-dependent enhancement of severe dengue disease in humans. Science, 358(6365):929–932, November 2017. ISSN 0036-8075, 1095-9203. doi: 10.1126/science.aan6836.

[21] Rosemary A. Aogo, Jose Victor Zambrana, Nery Sanchez, Sergio Ojeda, Guillermina Kuan, Angel Balmaseda, Aubree Gordon, Eva Harris, and Leah C. Katzelnick. Effects of boosting and waning in highly exposed populations on dengue epidemic dynamics. Science Translational Medicine, 15(722):eadi1734, November 2023. ISSN 1946-6234, 1946-6242. doi: 10.1126/scitranslmed.adi1734.

## References

[1] Aaron A. King, Dao Nguyen, and Edward L. Ionides. Statistical inference for partially observed Markov processes via the R package pomp. Journal of Statistical Software, 69(12): 1–43, 2016. doi: 10.18637/jss.v069.i12.

[2] Aaron A. King, Edward L. Ionides, Carles Martinez Bretó, Stephen P. Ellner, Matthew J. Ferrari, Sebastian Funk, Steven G. Johnson, Bruce E. Kendall, Michael Lavine, Dao Nguyen, Eamon B. O’Dea, Daniel C. Reuman, Helen Wearing, and Simon N. Wood. pomp: Statistical Inference for Partially Observed Markov Processes, 2024. URL https://kingaa.github.io/pomp/. R package, version 5.8.

[3] Zhilin Zeng, Juan Zhan, Liyuan Chen, Huilong Chen, and Sheng Cheng. Global, regional, and national dengue burden from 1990 to 2017: A systematic analysis based on the global burden of disease study 2017. EClinicalMedicine, 32, 2021.

